# Lost in Transplantation: Characterizing Racial Gaps in Physician Organ Offer Acceptance

**DOI:** 10.1101/2024.07.14.24310395

**Authors:** Hammaad Adam, Rene S. Bermea, Ming Ying Yang, Leo Anthony Celi, Marzyeh Ghassemi

## Abstract

**Background.:** There are known racial disparities in the organ transplant allocation system in the United States. However, prior work has yet to establish if transplant center decisions on offer acceptance—the final step in the allocation process— contribute to these disparities.

**Objective:** To estimate racial differences in the acceptance of organ offers by transplant center physicians on behalf of their patients.

**Design:** Retrospective cohort analysis using data from the Scientific Registry of Transplant Recipients (SRTR) on patients who received an offer for a heart, liver, or lung transplant between January 1, 2010 and December 31, 2020.

**Setting:** Nationwide, waitlist-based.

**Patients:** 32,268 heart transplant candidates, 102,823 liver candidates, and 25,780 lung candidates, all aged 18 or older.

**Measurements:** 1) Association between offer acceptance and two race-based variables: candidate race and donor-candidate race match; 2) association between offer rejection and time to patient mortality.

**Results:** Black race was associated with significantly lower odds of offer acceptance for livers (OR=0.93, CI: 0.88-0.98) and lungs (OR=0.80, CI: 0.73-0.87). Donor-candidate race match was associated with significantly higher odds of offer acceptance for hearts (OR=1.11, CI: 1.06-1.16), livers (OR=1.10, CI: 1.06-1.13), and lungs (OR=1.13, CI: 1.07-1.19). Rejecting an offer was associated with lower survival times for all three organs (heart hazard ratio=1.16, CI: 1.09-1.23; liver HR=1.74, CI: 1.66-1.82; lung HR=1.21, CI: 1.15-1.28).

**Limitations:** Our study analyzed the observational SRTR dataset, which has known limitations.

**Conclusion:** Offer acceptance decisions are associated with inequity in the organ allocation system. Our findings demonstrate the additional barriers that Black patients face in accessing organ transplants and demonstrate the need for standardized practice, continuous distribution policies, and better organ procurement.

## Introduction

Organ transplantation is a life-saving procedure for patients with end-stage diseases, but there is a severe shortage of transplant-viable organs from deceased donors.^1^ Prior work has established strong racial disparities in the system that coordinates the allocation of this scarce resource.^2–4^ For example, Black patients on the heart waitlist are less likely to receive a transplant and have a higher risk of post-transplant death.^3^ These inequities have persisted despite many changes to allocation policies, including the design of allocation scores such as the Model for End-Stage Liver Disease (MELD)^5^ and the Lung Allocation Score (LAS).^6^

In the US, the process to receive a transplant follows four steps (Fig. 1A).^7^ A patient is first referred by their physician to a transplant center (Referral), then evaluated by a multi-disciplinary committee for national waitlist placement (Transplant Evaluation and Committee Review). If approved, they are placed on the waitlist and assigned an allocation score, which determines the order in which donated organs are offered to waitlisted candidates (Waitlist Prioritization). When an organ is offered to a candidate, a physician at their transplant center decides to either accept or reject it on their behalf (Transplant Center Acceptance). Physicians assess the quality of the donated organ and its suitability for their patients using a wide range of information, including anthropometric data, lab values, and radiographic studies. If the offer is accepted, transplantation proceeds; otherwise, the organ is offered to the next-ranked candidate (Fig. 1B). Offer rejections are often made by physicians without input from patients and their families; we thus refer to these decisions as “physician-made.”

**Figure 1.**
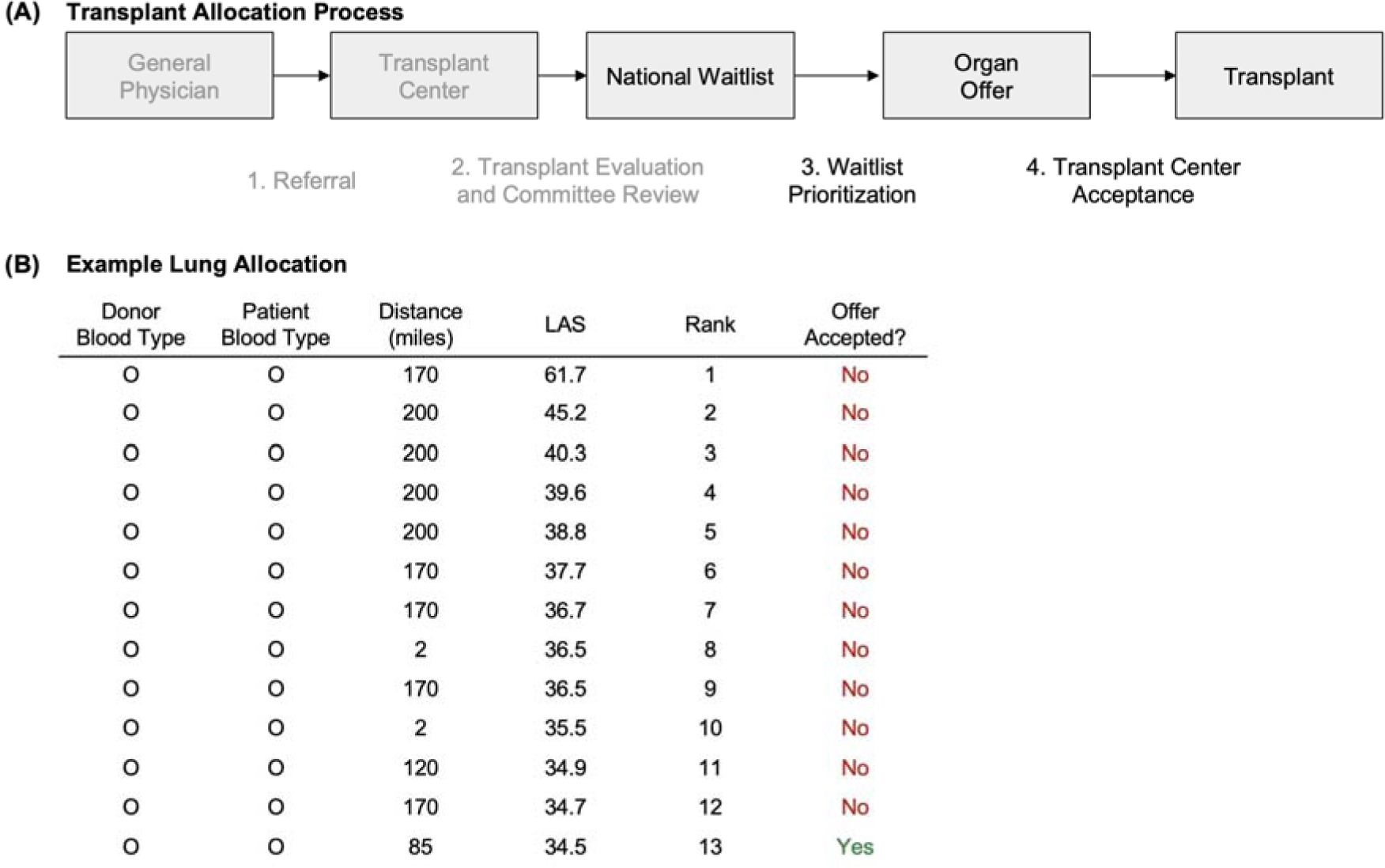
Overview of organ transplant allocation in the United States. A) Process to receive an organ transplant in the US. Patients are referred to a transplant center by a general physician. The transplant center evaluates the referral and if appropriate, convenes a multi-disciplinary clinical committee to determine candidacy for organ transplantation. If the patient is deemed to be a suitable transplant candidate, they are placed on a national waitlist. Medically compatible organs are offered to candidate recipients on the waitlist in order of a standardized allocation score or status. Once an offer is made, a transplant center physician decides whether to accept the organ on a candidate’s behalf. This final step means that top-ranked candidates do not always receive organs before lower-ranked candidates. Note that the first two steps are shown in gray, as data is only publicly available beyond this point. B) An example of lung allocation (from SRTR). The waitlist was ranked using blood type, distance between the donor and candidate, and the lung allocation score (LAS). All displayed candidates had the same blood type and distance bucket (< 250 miles), and so priority was determined by LAS alone. The thirteenth candidate on the waitlist received this organ for transplant, as physicians for the twelve higher-ranked candidates all rejected the offer.

While racial disparities in organ allocation are well-established, the equity impact of the final step–transplant center acceptance–is yet to be studied. These physician-made decisions are highly non-standardized: recent work has found large center-to-center variability in offer acceptance that directly impacts waitlist mortality.^8–11^ However, equity-focused changes to allocation policy – including the design of allocation scores^5,6^ – have largely focused on waitlist prioritization. Little work has examined whether transplant center acceptance decisions contribute to racial inequity in access to transplantation. This is a crucial gap in the literature, as no transplant can proceed without an offer being accepted.

In this study, we investigated the relationship between a candidate’s race and their transplant center’s decision to accept a heart, liver, or lung offer on their behalf. We estimated the association between offer acceptance and two race-based variables: candidate race and donor-candidate race match. We then estimated the association between increased offer rejection and patient survival, quantifying the cost of inequitable decision-making.

## Methods

### Data and Cohort Selection

This study used data from the Scientific Registry of Transplant Recipients (SRTR). The SRTR data system includes data on all donors, wait-listed candidates, and transplant recipients in the US, submitted by the members of the Organ Procurement and Transplantation Network (OPTN). The Health Resources and Services Administration (HRSA), U.S. Department of Health and Human Services provides oversight to the activities of the OPTN and SRTR contractors. The data reported here have been supplied by the Hennepin Healthcare Research Institute (HHRI) as the contractor for the Scientific Registry of Transplant Recipients (SRTR). The interpretation and reporting of these data are the responsibility of the author(s) and in no way should be seen as an official policy of or interpretation by the SRTR or the U.S. Government. This study was approved by the institutional review board at the Massachusetts Institute of Technology.

We conducted a retrospective cohort analysis using data on heart, liver, and lung transplant offers made between January 1, 2010 and December 31, 2020. These data are contained in SRTR’s Potential Transplant Recipients (PTR) files, which provide information on every offer made, including identifiers for the candidate and donor, candidate rank, whether the offer was accepted or rejected, and a reason for rejection. These data have been previously used in many studies.^8–10^ We merged these data with the SRTR standard analysis files, which provide key clinical and demographic information for each candidate and donor, including the allocation score—priority status for hearts, MELD for livers, and LAS for lungs—at the time of offer. Finally, we used candidate ZIP codes from SRTR and US government data sources to obtain two additional measures of candidate socio-demographics: ZIP-level household income (from the US Census^12^) and county-level social vulnerability index (from the Centers for Disease Control and Prevention^13^).

Figure 2 describes our cohort selection process. Our study exclusively considered offers that were rejected for one of two reasons: donor age or quality (OPTN refusal code 830) or donor size or weight (code 831). This exclusion is necessary as many rejection codes are driven by unobserved factors that are correlated with race. For example, human leukocyte antigen (HLA) types are correlated with race,^14^ but are not recorded in the data unless the candidate received a transplant. Including rejections for such reasons can lead to misleading results on the association between race and offer acceptance. In contrast, the included rejection codes— which comprise 73% of all rejections (Table S1)—are driven by observable factors that can be controlled for. This exclusion thus provides the most accurate estimates of the association between candidate race and offer acceptance. However, we emphasize that our main findings are unchanged if we relax this exclusion and include all rejection codes (see *Results*). Further, our exclusion criteria did not change the racial demographics of the cohort (Table S2).

**Figure 2.**
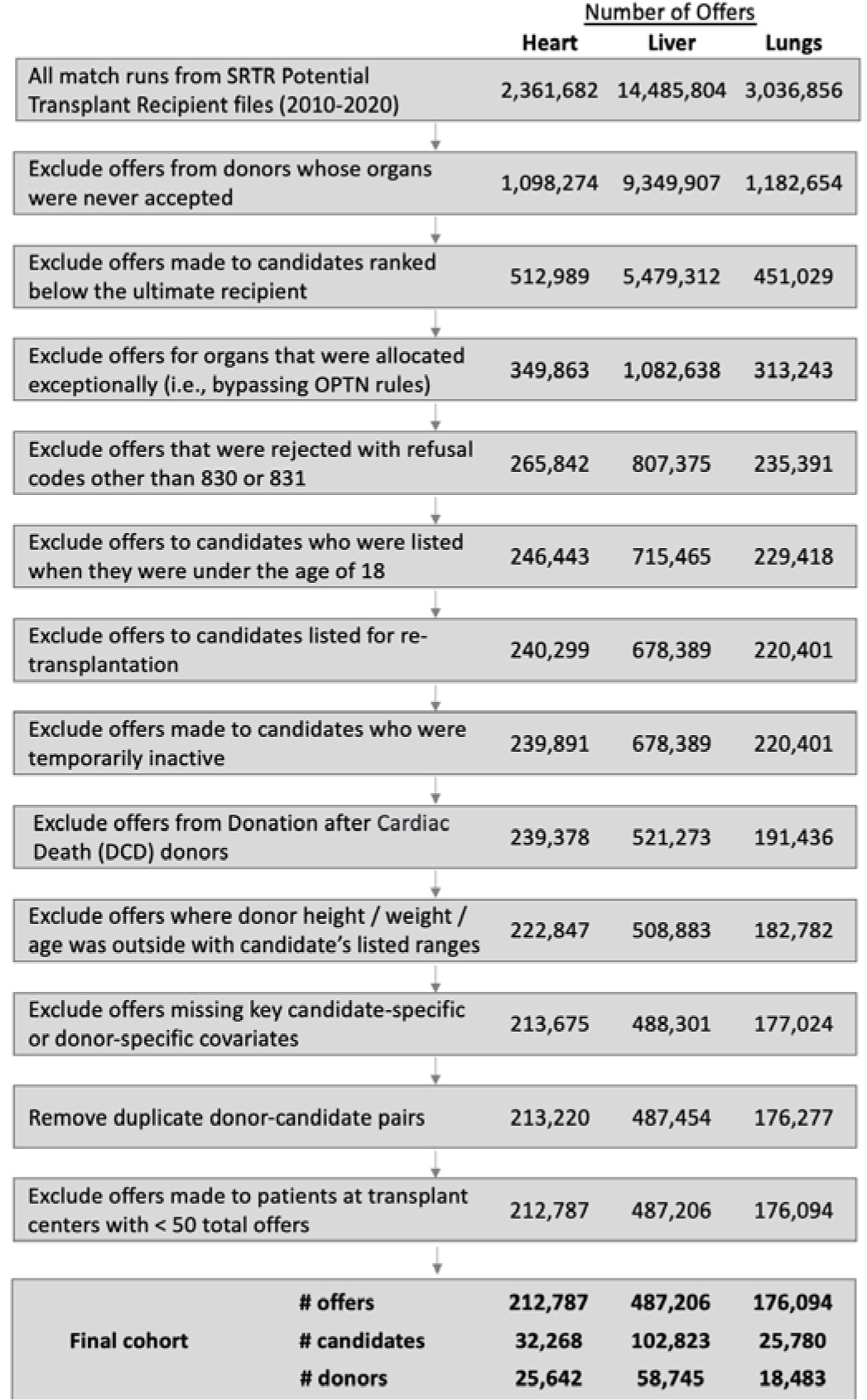
Cohort selection. We considered all offers made to patients on the waitlist for heart, liver, and lung transplants between January 1, 2010 and December 31, 2020. We excluded marginal organs that were not accepted by any candidate they were offered to. We excluded offers made to candidates ranked below the ultimate recipient in the waiting list, as these decisions were provisional and could be changed. We further excluded offers for organs that were allocated exceptionally (i.e. bypassing OPTN rules). To focus on decisions made based on observable characteristics and for non-logistical reasons, we only included rejected offers that were rejected for donor age or quality (code 830) or donor size or weight (code 831). We then excluded complex scenarios that required specialized decision-making and were not representative of a typical offer: this excluded candidates listed when they were under the age of 18, candidates listed for re-transplantation, heart candidates who were temporarily inactive, and organs donated after cardiac death (DCD). We did not include offers made to candidates where the donor did not meet their compatible age, weight, or height ranges (specified at the time of waitlisting), as these offers were rejected due to pre-specified criteria and did not require a physician decision at the time of offer. Finally, we excluded offers missing key candidate or donor covariates and offers made to patients at transplant centers with less than 50 total offers; this allowed us to adequately control for observed clinical features and center-level behavior respectively.

### Statistical Analysis

*Offer Acceptance.* We used multivariable logistic regressions to model the association between the binary outcome of offer acceptance and two race-based variables: 1) candidate race and 2) donor-candidate race match (i.e., race of the candidate and donor being the same). We encoded SRTR’s provider-reported race as a categorical variable with White as the reference level and Black or African-American, Hispanic or Latino, and Other as the non-reference levels. We used 80% of all offers in each organ cohort for model estimation and the remaining 20% for evaluation, generating this data split randomly. Recall that a donor organ may be offered to multiple candidates before it is accepted. Similarly, a candidate may reject multiple offers before they accept one and receive a transplant. Given this multiplicity, we calculated statistical significance using cluster-robust standard errors,^15^ clustering by candidate and donor. All analyses were conducted using R version 4.2.1.

We estimated two models for each organ type: partially adjusted and fully adjusted. The partially adjusted models controlled for a wide range of donor features (e.g., age, comorbidities, lab values; see Supplementary Methods for a full list). They included fixed effects for transplant center and year to control for mean center-level behavior and time trends respectively. They further controlled for the allocation score used for waitlist prioritization: priority status for hearts, MELD for livers, and LAS for lungs. The fully adjusted models controlled for several additional candidate characteristics, including clinical features (e.g., creatinine, functional status) and socio-demographics (e.g., ZIP-level household income). They also controlled for measures of candidate-donor compatibility such as the donor-candidate size ratio (measured using weight ratio for hearts and livers and predicted total lung capacity ratio for lungs). The covariates used for each organ type were chosen based on prior literature^2,8,10^ and clinical expertise. A subset of these covariates is summarized in Table 1 (see Supplementary Methods for the full list).

**Table 1A.** Candidate cohort demographics.

|  | **Heart**<br>**(N=32,268)** | **Liver**<br>**(N=102,823)** | **Lung**<br>**(N=25,780)** |
| --- | --- | --- | --- |
| **Age** |  |  |  |
| 18-35 | 3,098 (9.6%) | 5,239 (5.1%) | 2,405 (9.3%) |
| 36-50 | 6,738 (20.9%) | 17,026 (16.6%) | 3,183 (12.3%) |
| 51-65 | 15,931 (49.4%) | 59,099 (57.5%) | 12,175 (47.2%) |
| 66+ | 6,501 (20.1%) | 21,459 (20.9%) | 8,017 (31.1%) |
| **Race** |  |  |  |
| White | 20,553 (63.7%) | 71,947 (70.0%) | 20,358 (79.0%) |
| Black | 7,669 (23.8%) | 8,820 (8.6%) | 2,563 (9.9%) |
| Hispanic | 2,629 (8.1%) | 16,006 (15.6%) | 2,066 (8.0%) |
| Other | 1,417 (4.4%) | 6,050 (5.9%) | 793 (3.1%) |
| **Gender** |  |  |  |
| Female | 8,060 (25.0%) | 37,318 (36.3%) | 11,287 (43.8%) |
| Male | 24,208 (75.0%) | 65,505 (63.7%) | 14,493 (56.2%) |
| **Blood Type** |  |  |  |
| A | 12,184 (37.8%) | 38,230 (37.2%) | 10,022 (38.9%) |
| AB | 1,587 (4.9%) | 4,148 (4.0%) | 947 (3.7%) |
| B | 4,636 (14.4%) | 12,359 (12.0%) | 2,896 (11.2%) |
| O | 13,861 (43.0%) | 48,086 (46.8%) | 11,915 (46.2%) |
| **BMI** |  |  |  |
| 0-18.5 | 648 (2.0%) | 1,315 (1.3%) | 1,722 (6.7%) |
| 18.5-24.9 | 8,272 (25.6%) | 22,288 (21.7%) | 8,494 (32.9%) |
| 25-29.9 | 11,635 (36.1%) | 35,593 (34.6%) | 9,698 (37.6%) |
| 30+ | 11,713 (36.3%) | 43,627 (42.4%) | 5,866 (22.8%) |
| **Insurance Type** |  |  |  |
| Public | 15,593 (48.3%) | 45,540 (44.3%) | 12,846 (49.8%) |
| Private | 16,497 (51.1%) | 56,502 (55.0%) | 12,798 (49.6%) |
| Other | 178 (0.6%) | 781 (0.8%) | 136 (0.5%) |

|  | Heart<br>(N=32,268) | Liver<br>(N=102,823) | Lung<br>(N=25,780) |
| --- | --- | --- | --- |
| **County-level Social Vulnerability Index** |  |  |  |
| Mean (SD) | 0.477 (0.215) | 0.484 (0.215) | 0.446 (0.207) |
| **ZIP-level Median Household Income** |  |  |  |
| Mean (SD) | 68,600 (27,700) | 68,500 (27,300) | 70,500 (27,600) |
| **Primary Diagnosis Group** |  |  |  |
| CAD | 806 (2.5%) | 0 (0%) | 0 (0%) |
| Cardiomyopathy | 29,928 (92.7%) | 0 (0%) | 0 (0%) |
| Congenital | 1,093 (3.4%) | 0 (0%) | 0 (0%) |
| Other | 19 (0.1%) | 4,091 (4.0%) | 28 (0.1%) |
| Valve Replacement/Repair | 422 (1.3%) | 0 (0%) | 0 (0%) |
| Acute Hepatic Necrosis | 0 (0%) | 3,020 (2.9%) | 0 (0%) |
| Alcoholic Liver Disease/Cirrhosis | 0 (0%) | 27,995 (27.2%) | 0 (0%) |
| Biliary Atresia | 0 (0%) | 245 (0.2%) | 0 (0%) |
| Cholestatic Liver Disease/Cirrhosis | 0 (0%) | 6,977 (6.8%) | 0 (0%) |
| Malignant Neoplasms | 0 (0%) | 11,684 (11.4%) | 0 (0%) |
| Metabolic Disease | 0 (0%) | 1,801 (1.8%) | 0 (0%) |
| Non-cholestatic and Non-alcoholic Cirrhosis | 0 (0%) | 47,010 (45.7%) | 0 (0%) |
| Cystic Fibros | 0 (0%) | 0 (0%) | 2,620 (10.2%) |
| Obstructive | 0 (0%) | 0 (0%) | 7,297 (28.3%) |
| Restrictive | 0 (0%) | 0 (0%) | 14,487 (56.2%) |
| Vascular | 0 (0%) | 0 (0%) | 1,348 (5.2%) |
| **Heart Candidate Priority Status** |  |  |  |
| Status 1A | 22,295 (69.1%) |  |  |
| Status 1B | 8,518 (26.4%) |  |  |
| Status 2 | 1,455 (4.5%) |  |  |
| **MELD Score** |  |  |  |
| Mean (SD) |  | 25.7 (9.15) |  |
| **Calculated LAS score** |  |  |  |
| Mean (SD) |  |  | 47.9 (18.1) |

<sup>1</sup> Note that only a small percentage of heart patients were listed under the new 6-tiered prioritization system (Status 1-6), so all new priority statuses were mapped to the prior 3-tiered prioritization system (Status 1A, 1B, and 2) for analysis.

**Table 1B.** Donor cohort demographics.

|  | **Heart**<br>**(N=25,642)** | **Liver**<br>**(N=58,745)** | **Lung**<br>**(N=18,483)** |
| --- | --- | --- | --- |
| **Age** |  |  |  |
| 18-35 | 15,787 (61.6%) | 22,008 (37.5%) | 9,966 (53.9%) |
| 36-50 | 7,768 (30.3%) | 15,873 (27.0%) | 5,028 (27.2%) |
| 51-65 | 2,080 (8.1%) | 15,925 (27.1%) | 3,233 (17.5%) |
| 66+ | 7 (0.0%) | 4,939 (8.4%) | 256 (1.4%) |
| **Race** |  |  |  |
| White | 16,179 (63.1%) | 37,431 (63.7%) | 11,170 (60.4%) |
| Black | 4,284 (16.7%) | 11,404 (19.4%) | 3,615 (19.6%) |
| Hispanic | 4,395 (17.1%) | 7,820 (13.3%) | 2,928 (15.8%) |
| Other | 784 (3.1%) | 2,090 (3.6%) | 770 (4.2%) |
| **Gender** |  |  |  |
| Female | 7,745 (30.2%) | 23,875 (40.6%) | 7,386 (40.0%) |
| Male | 17,897 (69.8%) | 34,870 (59.4%) | 11,097 (60.0%) |
| **Blood Type** |  |  |  |
| A | 9,234 (36.0%) | 21,876 (37.2%) | 6,673 (36.1%) |
| AB | 573 (2.2%) | 1,955 (3.3%) | 436 (2.4%) |
| B | 2,872 (11.2%) | 7,292 (12.4%) | 2,103 (11.4%) |
| O | 12,963 (50.6%) | 27,622 (47.0%) | 9,271 (50.2%) |
| **Death Mechanism** |  |  |  |
| Asphyxiation | 1,514 (5.9%) | 2,541 (4.3%) | 858 (4.6%) |
| Blunt Injury | 7,340 (28.6%) | 11,249 (19.1%) | 4,271 (23.1%) |
| Cardiovascular | 2,190 (8.5%) | 9,109 (15.5%) | 1,450 (7.8%) |
| Drug Intoxication | 3,780 (14.7%) | 6,895 (11.7%) | 1,927 (10.4%) |
| Intracranial Hemorrhage/Stroke | 4,725 (18.4%) | 19,694 (33.5%) | 5,709 (30.9%) |
| Other | 6,093 (23.8%) | 9,257 (15.8%) | 4,268 (23.1%) |
| **Did the Donor Meet CDC Guidelines for High Risk?** |  |  |  |
| No | 19,271 (75.2%) | 45,496 (77.4%) | 14,727 (79.7%) |
| Unknown | 4 (0.0%) | 9 (0.0%) | 4 (0.0%) |
| Yes | 6,367 (24.8%) | 13,240 (22.5%) | 3,752 (20.3%) |
| **Smoking History** |  |  |  |
| No | 22,438 (87.5%) | 44,925 (76.5%) | 16,847 (91.1%) |
| Unknown | 265 (1.0%) | 850 (1.4%) | 189 (1.0%) |
| Yes | 2,939 (11.5%) | 12,970 (22.1%) | 1,447 (7.8%) |

The difference between the partially and fully adjusted models is the included set of candidate covariates. While the partially adjusted models controlled only for waitlist priority, the fully adjusted models controlled for all candidate covariates. A significant partially-adjusted association between race and offer acceptance indicates that candidates of a given race are less likely to have offers accepted than candidates of another race with the same waitlist priority (conditional on donor quality, year, and transplant center). This association may be explained by other candidate features (e.g., creatinine); however, it still reflects an inequity, as it indicates a barrier to transplant access that is not balanced by additional waitlist priority. Meanwhile, a fully-adjusted association indicates a racial discrepancy that cannot be explained by any observed feature of the donor or candidate.

*Patient Outcomes*. In addition to modeling offer acceptance, we used a subset of our cohort to estimate the impact of rejecting an offer on patient survival. We identified pairs of candidates such that the two candidates were offered the same organ, one candidate accepted the offer while the other candidate rejected the offer, and the rejecting candidate was ranked immediately above the accepting candidate. This approach yielded a paired dataset where the accepting and rejecting candidates for each donor organ were well-matched on waitlist priority and illness severity (see Table S3). To prevent any candidate from appearing in both the accepting and rejecting groups, we excluded pairs where the accepting patient had previously rejected an offer. We then compared outcomes between the accepting and rejecting groups using Cox regressions with offer rejection as the exposure and mortality as the endpoint. These regressions were stratified by donor to account for the paired nature of the data, and controlled for time on the waitlist in addition to all candidate covariates described previously. Confidence intervals for all models were calculated using robust standard errors. To protect against proportional hazards violations, we also estimated differences in restricted mean survival time (RMST).^16^

## Results

Our offer acceptance cohort consisted of 212,787 heart transplant offers made to 32,268 candidates, 487,206 liver transplant offers to 102,823 candidates, and 176,094 lung transplant offers to 25,780 candidates. Table 1 summarizes the demographics of candidates and donors in our cohort. A simple descriptive analysis indicates that a donor organ was usually rejected by several physicians before being accepted for its final recipient (Fig. 3A). A candidate typically received several offers before they had one accepted (or were removed from the waitlist due to e.g., death) (Fig. 3B). There was a clear discrepancy between the racial demographics of first-ranked candidates and eventual transplant recipients (Fig. 3C). The results of our full statistical analysis are displayed in Figure 4. We focus on four key findings.

**Figure 3.**
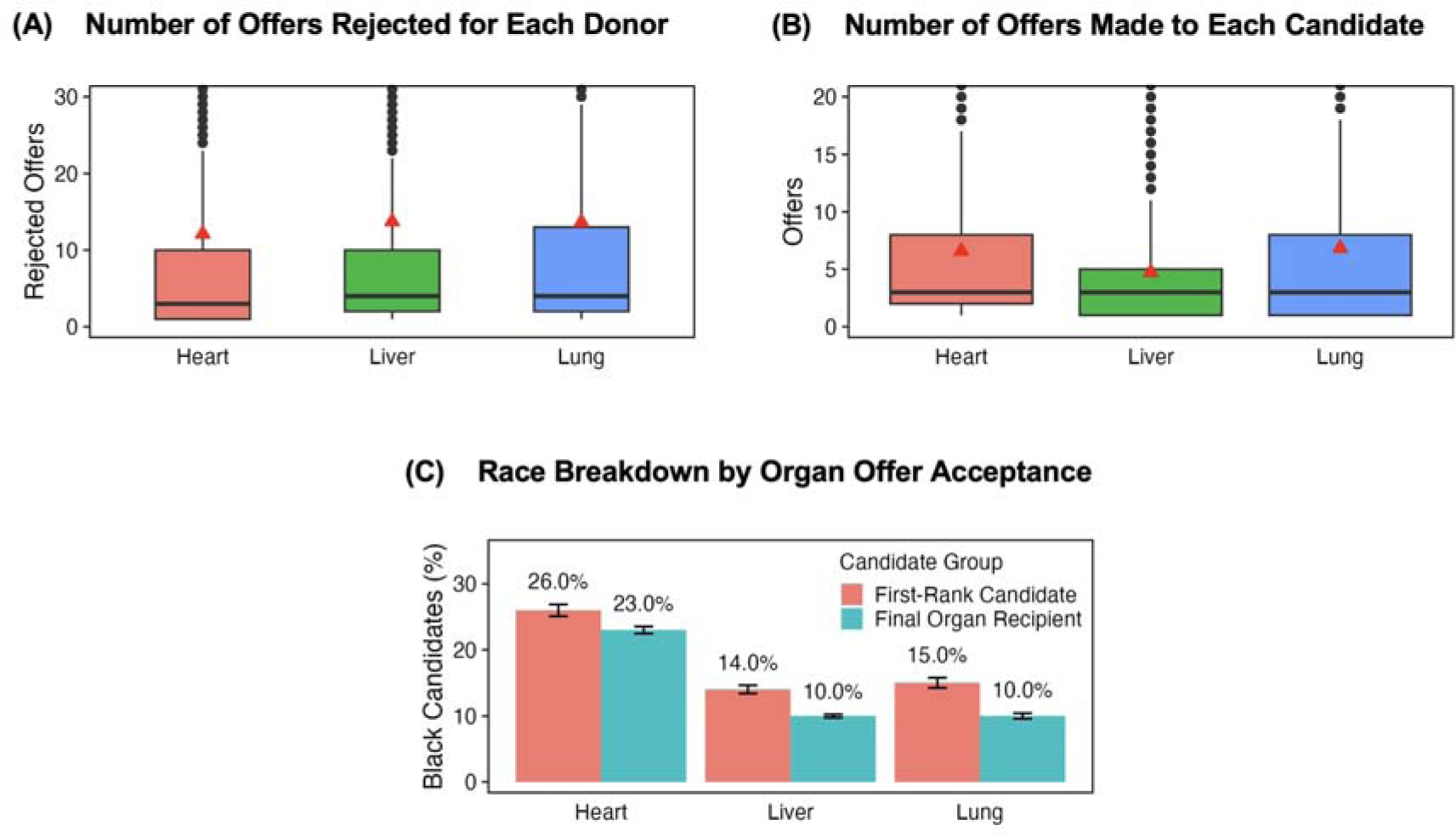
Organ offer acceptance patterns. A) Distribution of the number of offers rejected before an organ is accepted for its final recipient. The plot displays the mean (red triangle), median (black line), and interquartile range (box) of the number of offers rejected. On average, between eleven and fifteen candidates (depending on the organ type) reject an organ before it is accepted for its recipient. B) Distribution of the number of offers received by each candidate in our cohort. The plot displays the mean (red triangle), median (black line), and interquartile range (box). On average, a candidate in our cohort receives between five and seven offers before they either accept (and receive a transplant) or are removed from the waitlist (e.g., due to death). C) Racial disparities between final organ recipients and first-ranked candidates for each donated organ. A significantly higher proportion of first-ranked candidates are Black compared to actual recipients for liver and lung transplants (p < 0.001, chi-squared test). The error bars denote the 95% confidence intervals of the proportion (calculated using the normal approximation to the binomial distribution).

**Figure 4.**
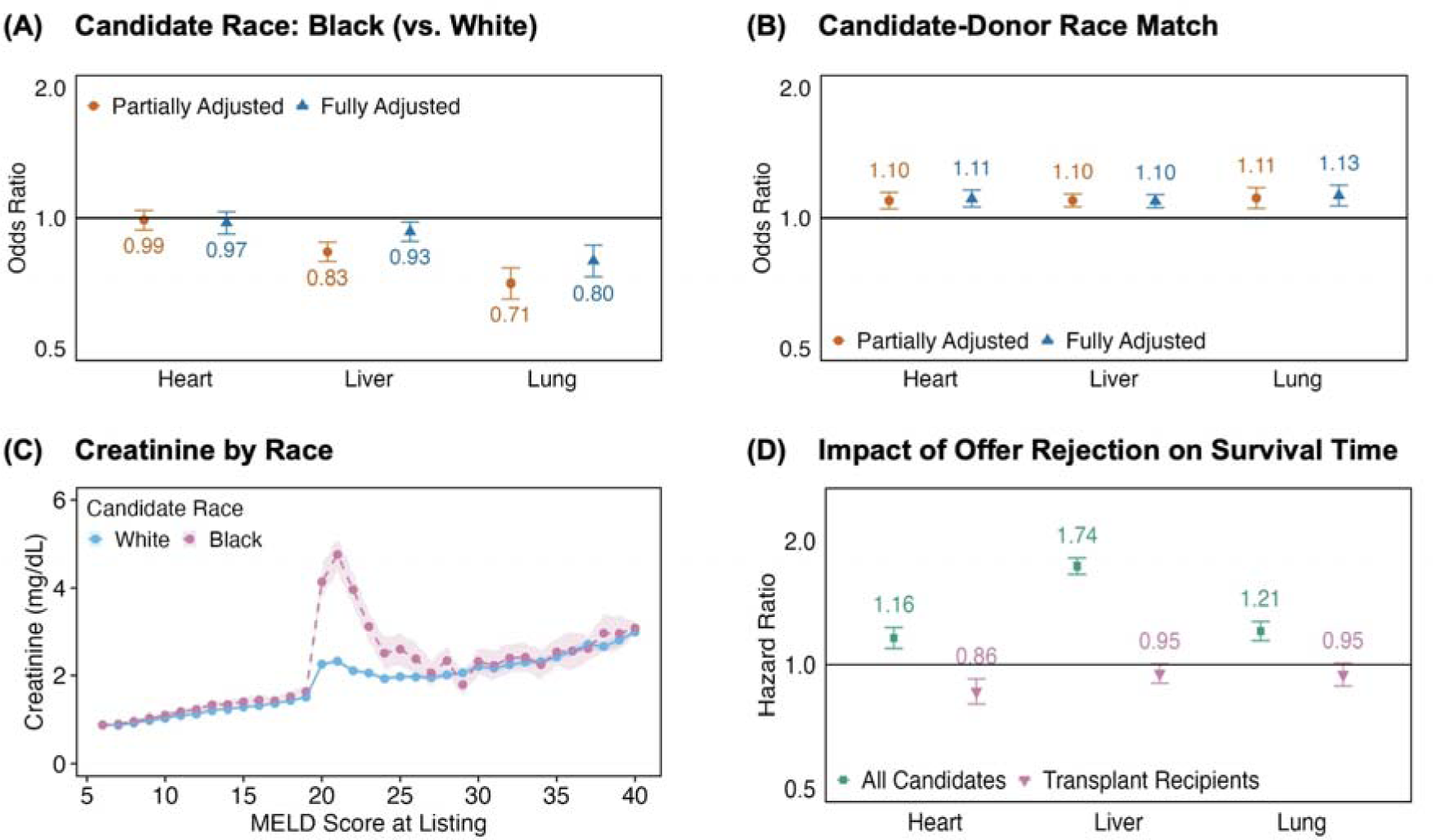
Modeling organ transplant offer acceptance and patient survival. A) Odds ratios for the association between Black candidate race and offer acceptance. Black race is associated with significantly lower liver and lung offer acceptance in the partially adjusted and fully adjusted models. We plot odds ratios (on a log-scale) from multivariable logistic regressions modeling offer acceptance. Error bars denote 95% confidence intervals derived from cluster-robust standard errors (clustering by candidate and donor). B) Odds ratios for the association between donor-candidate race match and offer acceptance. A donor-candidate race match is associated with significantly higher odds of offer acceptance for all three organ types. We plot odds ratios (on a log-scale) from multivariable logistic regressions modeling offer acceptance. Error bars denote 95% confidence intervals, calculated using cluster-robust standard errors (clustering by candidate and donor). C) Mean serum creatinine levels by MELD score at listing for Black and White candidates in our liver cohort. On average, Black patients have higher levels of creatinine than White patients with the same MELD score, particularly in the 19-24 point range. The shaded bands display 95% confidence intervals of the mean. D) Hazard ratios for the impact of offer rejection on time to mortality. Candidates with rejected offers have increased mortality risk than similarly ill candidates with accepted offers (“All Candidates” group). Even if the rejecting candidate ultimately received a transplant (“Transplant Recipients” group), they did not receive significantly higher survival benefit from the future accepted organ than the present rejected organ. We plot hazard ratios (on a log-scale) from stratified Cox proportional hazards models. Error bars denote 95% confidence intervals calculated using robust standard errors.

### Physicians were less likely to accept liver and lung offers on behalf of Black candidates

We first consider our models of offer acceptance. We confirmed that these models fit the data well: they were highly predictive and well-calibrated when evaluated on the held-out set of offers (Fig. S1). We focus specifically on the estimated association between Black candidate race (vs. White race) and a physician’s decision to accept an organ offer (Fig. 4A). We found that Black race was associated with significantly lower odds of lung offer acceptance, both in the partially adjusted (odds ratio=0.71, 95% confidence interval: 0.65-0.77) and fully adjusted (OR=0.80, 95% CI: 0.73-0.87) cases. We observed a similar pattern for livers, though with smaller disparities (partially adjusted OR=0.83, 95% CI: 0.79-0.88, fully adjusted OR=0.93, CI: 0.88-0.98). We observe weaker evidence of disparities against Hispanic patients in the partially adjusted models for livers and lungs (Fig. S2). Full regression coefficients for all models are included in the Supplement (Tables S4-S6).

We briefly discuss potential drivers and modifiers of these results. We confirm that the results are not driven by our rejection code exclusion criterion; our main findings are unchanged if we include fewer, more, or all rejection codes (Fig. S3). The observed associations for Black race were not explained by differences in observed covariates, including blood type, diagnosis, illness severity, county socioeconomics, or size mismatches. The associations did not vary by transplant center size (Table S7). The observed discrepancies have worsened over time (Table S8): the odds ratios for Black race for liver and lung offers were significantly lower in the last five years (2016-2020) in our cohort than the first six years (2010-2015). While Black race was not associated with reduced heart offer acceptance, an analysis of the fully-adjusted fixed effects found that Black candidates were over-represented at transplant centers with lower mean acceptance propensities (Fig. S4). No such association was observed for livers and lungs.

### MELD and LAS did not sufficiently account for racial differences in clinical characteristics

For liver and lung transplants, the significant effects of Black race in the partially adjusted models imply that physicians were less likely to accept offers for Black candidates than White candidates with the same waitlist priority (i.e., MELD or LAS). These coefficients were reduced in the fully adjusted models, implying that there exist candidate characteristics that were correlated with race and impacted offer acceptance, but were not fully captured by MELD and LAS. For example, we found that high creatinine–a marker of renal dysfunction^17^–was associated with lower offer acceptance (OR=0.95, CI: 0.94 - 0.96) and that Black patients in our cohort had higher levels of creatinine than White patients with the same MELD (Fig. 4C). Note that while the MELD equation includes creatinine, it caps its value at 4 mg/dl.^18^ This cutoff was exceeded by 13% of Black patients and 6% of White patients in our liver cohort; MELD may thus not adequately capture renal function for these patients.

### Race concordance increased offer acceptance for hearts, livers, and lungs

We found that a donor-candidate race match was associated with significantly higher odds of offer acceptance for hearts (OR=1.11, 95% CI: 1.06-1.16), livers (OR=1.10, 95% CI: 1.06-1.13), and lungs (OR=1.13, 95% CI: 1.07-1.19) (Fig. 4B). As before, this behavior was not explained by racial differences in the observed covariates, including blood type, size, or other clinical characteristics. They were also not explained by racial differences in HLA types, since we excluded rejections due to HLA mismatches. A preference for racially concordant transplants can disadvantage Black patients due to the relatively fewer number of Black organ donors. For example, Black patients made up 24% of waitlisted candidates but only 17% of donors in our heart cohort (Table 1).

### Offer rejection was associated with worse patient outcomes

Finally, we consider the impact of offer rejection on downstream patient outcomes. Our outcomes cohort consisted of 13,546 heart candidates, 27,370 liver candidates, and 11,184 lung candidates. Cox regressions (Fig. 4D, “All Candidates”) indicated that offer rejection was associated with significantly lower survival times for hearts (hazard ratio=1.16, 95% CI: 1.09-1.23), livers (HR=1.74, 95% CI: 1.66-1.82), and lungs (HR=1.21, 95% CI: 1.15-1.28).

This association was largely driven by the risk of pre-transplant mortality: for example, 16% of rejecting liver candidates died or deteriorated before they could accept another offer. To isolate the risk of pre-transplant mortality from the quality of the organ received, we further narrowed our cohort to only include donor organs for which the rejecting candidate later received a different transplant. This approach ignores the risk of pre-transplant death and compares the benefit of the future accepted transplant to the present rejected one. Here, offer rejection was not significantly associated with survival (Fig. 4D, “Transplant Recipients”) for livers and lungs, though it was associated with increased survival for hearts (HR=0.86, 95% CI 0.80-0.92). RMST-based analyses provided similar results (Fig. S5).

## Discussion

Black race was associated with significantly lower odds of offer acceptance for liver and lung transplants. This pattern was particularly concerning for lungs, where Black patients had 20% lower odds of offer acceptance than White patients with similar clinical characteristics. Only one existing paper has studied the association between offer acceptance and race. This article^19^—which exclusively studied heart offers—found that the cumulative incidence of heart offer acceptance was significantly lower for Black candidates. Our work builds upon this analysis by illustrating that similar patterns exist for lungs and livers and by highlighting the downstream impact of offer rejection on patient survival. Note that our analysis did not uncover a similar association for hearts, likely due to the different time period considered. The observed associations between race and offer acceptance are concerning, especially as prior work has found similar associations between race and other key decisions made by transplant centers.^20,21^ Overall, our findings highlight the need for greater transparency and standardization in transplant center decision-making.

It is possible that the observed racial discrepancies were not due to physician bias, but rather explained by unobserved candidate characteristics. However, our analysis demonstrates that the observed associations reflect inequities in the allocation system even if they were driven by unbiased decision-making. We found that allocation scores such as MELD and LAS did not adequately account for observed variables that impact offer acceptance. Consider the example of serum creatinine in liver transplants. We found that conditional on MELD, high-creatinine patients were more likely to have offers rejected than low-creatinine patients. This decision-making may be clinically justified, as renal dysfunction is a risk factor for poor outcomes following transplantation.^22^ However, Black patients were more likely to have high creatinine than White patients with the same waitlist priority (i.e., MELD); they thus needed more offers (on average) to find suitable livers, as each offer was more likely to be rejected.

Allocation scores not accounting for offer acceptance patterns puts Black patients at a systemic disadvantage: it is harder to find them suitable organs, but they receive no additional priority in the waitlist to overcome this obstacle. Thus, under current waitlist design, even unbiased physician decision-making can create inequity. This shortcoming can be addressed by increasing waitlist priority for patients who are medically harder-to-match. OPTN recently took a step in this direction for lung transplantation by adopting continuous distribution, a policy that prioritizes candidates not only based on medical urgency and transplant benefit, but also hardness-to-match, patient access, and logistical efficiency.^23^ As part of this policy shift, the Lung Allocation Score was replaced by the Continuous Allocation Score (CAS) in March 2023.^24^ While it is too early to evaluate the impact of CAS on lung allocation, our findings suggest that this shift can have a positive impact on equity.

Our study further revealed a physician preference for race-matched transplant allocations that could not be explained by racial differences in blood type, size, or other clinical features. We found that a donor-candidate race match increased the odds of physician offer acceptance by 10-13%. This preference is concerning, as there is only mixed evidence on the benefits of a race match on long-term transplant outcomes.^25–28^ Even if such a preference is clinically justified, it is concerning given historical inequities in organ procurement. Organ procurement organizations (OPOs)–the nonprofits that are tasked with procuring organs from deceased donors–have historically failed to procure a large number of organs from Black and other non-White donors.^29^ Our analysis thus suggests that any changes in organ allocation systems can only go so far: to ensure equity, organ procurement must improve too.

Finally, our study clarifies the cost of offer rejection. Physicians may reject an organ on behalf of their patient if they believe they may receive a better offer in the future. However, we found that such decisions greatly increased the risk of death in the interim. Further, for livers and lungs, even if the candidate later received a transplant, the accepted organ did not provide significantly higher survival benefit than the rejected organ. These results imply that waiting is unlikely to be worth the added mortality risk. Black patients were more likely to bear this cost, as our analysis establishes that they were more likely to have liver and lung offers rejected. Our study thus suggests that physician decision-making on transplant offers contributes to previously documented racial gaps in waitlist mortality.^3,4^

Our work has several limitations. First, to minimize the effect of race-correlated unobservables, our main analysis focused only on two rejection codes. However, the codes noted in the SRTR data may not always reflect the true reasons for rejection; for example, “donor age or quality” may be used as a catch-all category for a long tail of reasons. We emphasize that our conclusions are robust to this limitation: our key findings remain unchanged if we analyze fewer, more, or all rejection codes. Second, the SRTR data does not provide granular socioeconomic measures beyond insurance type. While we controlled for median measures in the candidate’s zip code or county of residence, this approach is unlikely to capture the full scope of a candidate’s social support. Third, our analysis did not focus on kidney transplantation. Not only are kidneys the most transplanted organ, but also their allocation system has been particularly prone to racial inequities.^30,31^ However, unlike the three organs considered, race may explicitly be involved in kidney offer acceptance. Several scores used in kidney transplantation—including the kidney donor risk index^32^ and eGFR^33^ (during our study period)—explicitly adjust for Black race. A similar analysis of kidney offers would need to account for the association between race and rejection that stems solely from the widespread use of these scores. We thus did not include kidney transplantation in this study, as this nuance merits careful consideration in a separate manuscript.

In conclusion, our study demonstrates the additional barriers Black patients face in transplant access due to transplant center decision-making. To address the identified inequities, we advocate for greater transparency and standardization in acceptance decisions, well-designed continuous distribution policies, and a strong policy focus on equitable organ procurement.

## Data Availability

This study used data from the Scientific Registry of Transplant Recipients (SRTR).

## Acknowledgments

We thank Irene Chen, Matthew McDermott, Emily Alsentzer, Tom Hartvigsen, and Aparna Balagopalan for their helpful comments. This work was supported by funding from the MIT Jameel Clinic. This research was supported, in part, by Takeda Development Center Americas, INC. (successor in interest to Millennium Pharmaceuticals, INC.). RSB is supported by an NIH Ruth L. Kirschstein National Research Service Award (T32HL116275). MG is supported in part by a CIFAR AI Chair at the Vector Institute. LAC is funded by the National Institute of Health through the NIBIB R01 grant EB017205.

## Authors’ contributions

Conceptualization: HA, RSB, LAC, MG

Methodology: HA, MY

Formal analysis, investigation, and visualization: HA, MY

Funding acquisition: LAC, MG

Supervision: LAC, MG

Writing – original draft: HA, MY, RSB

Writing – review & editing: HA, RSB, MY, LAC, MG

## Conflicts of interest

Authors declare that they have no conflicts of interests.

## Role of funding source

The funders of the study had no role in study design, data collection, data analysis, data interpretation, or writing of the article.

## Ethics Committee approval

This study was granted full approval following expedited review by COUHES, the Institutional Review Board for the Massachusetts Institute of Technology.

## Supplementary Materials

### Supplementary Methods

#### Full List of Covariates

The partially adjusted models controlled for the following donor features: age, gender, race, blood type, smoking history, death mechanism, diabetes, presence of infection, and high risk status (as defined by the Centre for Disease Control and Prevention). We additionally controlled for bilirubin, blood urea nitrogen (BUN), creatinine, cocaine use, and ejection fraction for heart donors, bilirubin and creatinine for liver donors, and partial pressure of oxygen (PO2), abnormal chest X-ray, and bronchoscopy results for lung donors. On the candidate side, the partially adjusted models controlled only for the allocation score used in waitlist prioritization: MELD for livers, LAS for lungs, and priority status for hearts. Only a small percentage of heart patients were listed under the new 6-tiered prioritization system (Status 1-6), so all new priority statuses were mapped to the prior 3-tiered prioritization system (Status 1A, 1B, and 2)^35^ for analysis. Broadly, these allocation scores capture how urgently a patient needs a transplant, and dictate the order in which donated organs are offered to potential recipients. Moreover, in line with prior research^10^, we controlled for PO2, LAS, and MELD using a restricted cubic spline with four knots.

In addition to these features, the fully adjusted models controlled for a comprehensive set of candidate variables that capture clinical conditions, social determinants, and donor-specific compatibility. We included fixed effects for a candidate’s transplant center and the year of offer to account for previously described variability in transplant center decision-making^8–10^ and time trends. We further controlled for the following candidate features: age, gender, blood type, primary diagnosis, insurance, preliminary crossmatch requirement, median income and social vulnerability in residence zip code, body mass index (BMI), diabetes status, functional status, dialysis use, extracorporeal membrane oxygenation (ECMO) use and mechanical ventilation use. We also include organ-specific candidate information: creatinine, previous surgery type, Ventricular Assist Device type, intra-aortic balloon pump (IABP) use, intravenous inotropic medication use, number of prior sternotomies, and smoking history for heart candidates, albumin, sodium, creatinine, bilirubin, and history of angina or coronary artery disease for liver candidates, and transplant type (single vs bilateral), smoking history, and mean pulmonary artery pressure (as a restricted cubic spline with four knots) for lung candidates. Finally, we included a set of features that capture specific elements of donor-candidate compatibility: gender match, blood type match (identical vs. non-identical), and organ size match. We accounted for organ size match by controlling for predicted total lung capacity (pTLC) ratio for lungs (as a restricted cubic spline with four knots) and weight ratio for liver and hearts (as a restricted cubic spline with three knots).

### Supplementary Figures

**Figure S1.**
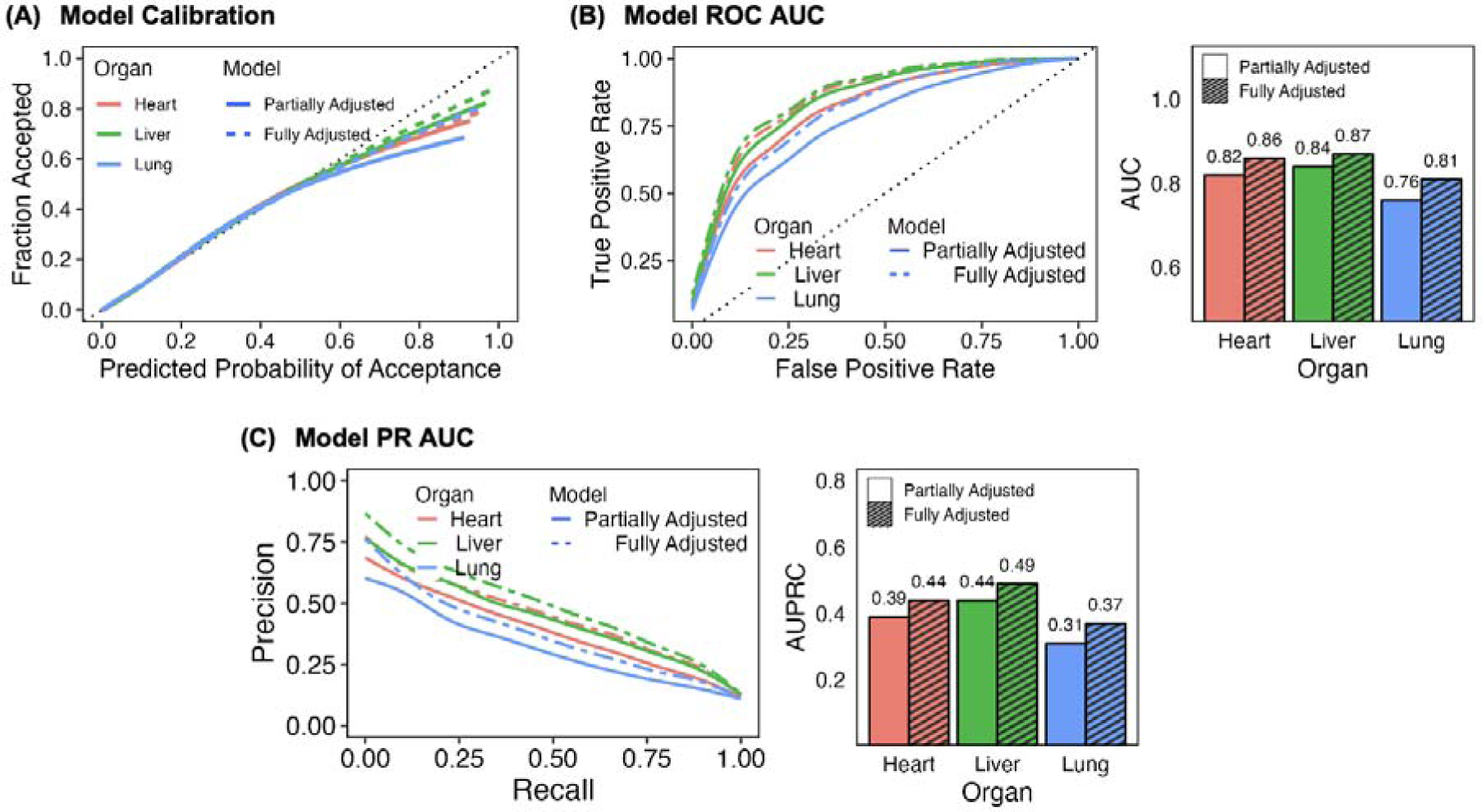
Offer acceptance model evaluation. A) Calibration curves for the logistic regressions modeling offer acceptance. All models are well-calibrated. The calibration curve for each model and organ type is smoothed (using a generalized additive model) and plotted. B) Predictive accuracy of the logistic regressions used to model offer acceptance for heart, liver, and lung transplants. Accuracy is measured using the area under the receiver operating characteristic curve (ROC AUC). All models have high predictive accuracy on held-out data, with ROC AUCs exceeding 0.75 for all three organ types. The fully adjusted models are particularly accurate, with ROC AUCs over 0.8. C) Trade-off between precision and recall. The area under the precision-recall curve (PR AUC) is commonly used to evaluate model performance for imbalanced data. All models have PR AUCs far greater than 0.11, which is approximately the proportion of positive labels (offer acceptance) in the dataset.

**Figure S2.**
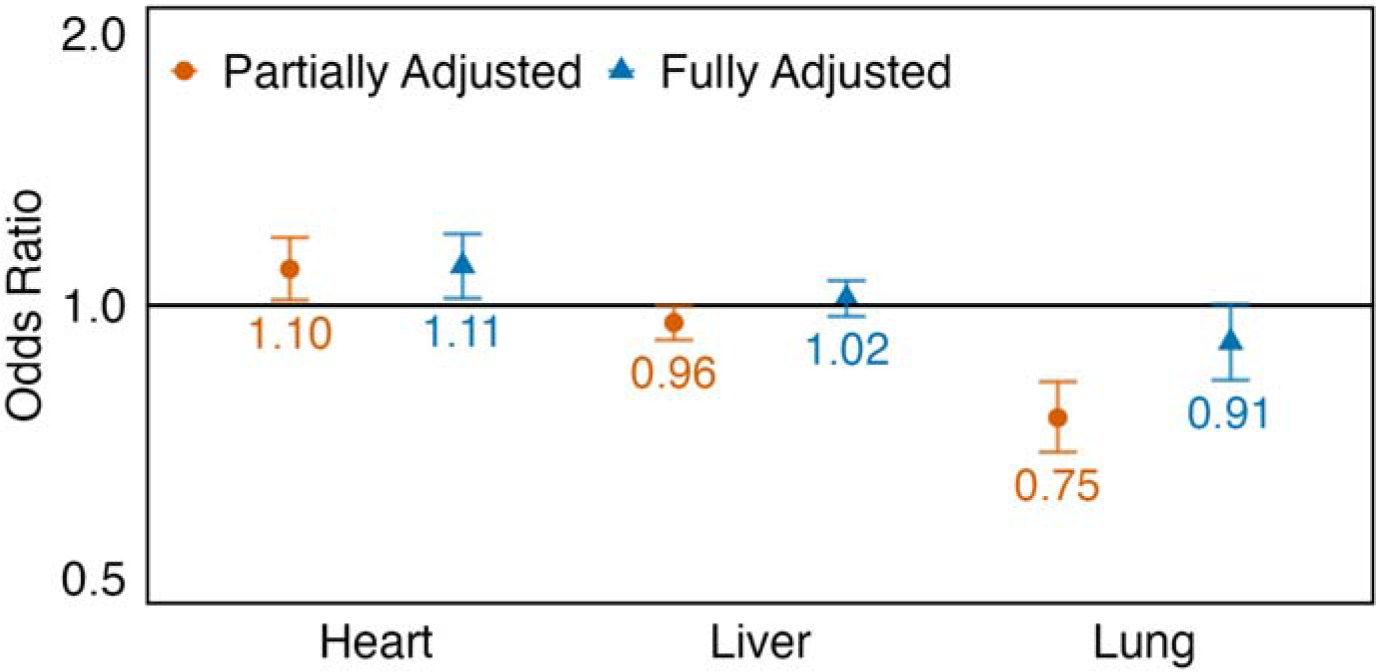
Organ offer acceptance: effect of Hispanic (vs. White) race. Odds ratios for the association between Hispanic candidate race (vs. White race) and offer acceptance. Hispanic patient race is associated with significantly lower liver and lung offer acceptance in the partially adjusted models. Hispanic race is also associated with significantly higher offer acceptance in both heart models. Error bars denote 95% confidence intervals, calculated using cluster-robust standard errors (clustering by candidate and donor).

**Fig S3.**
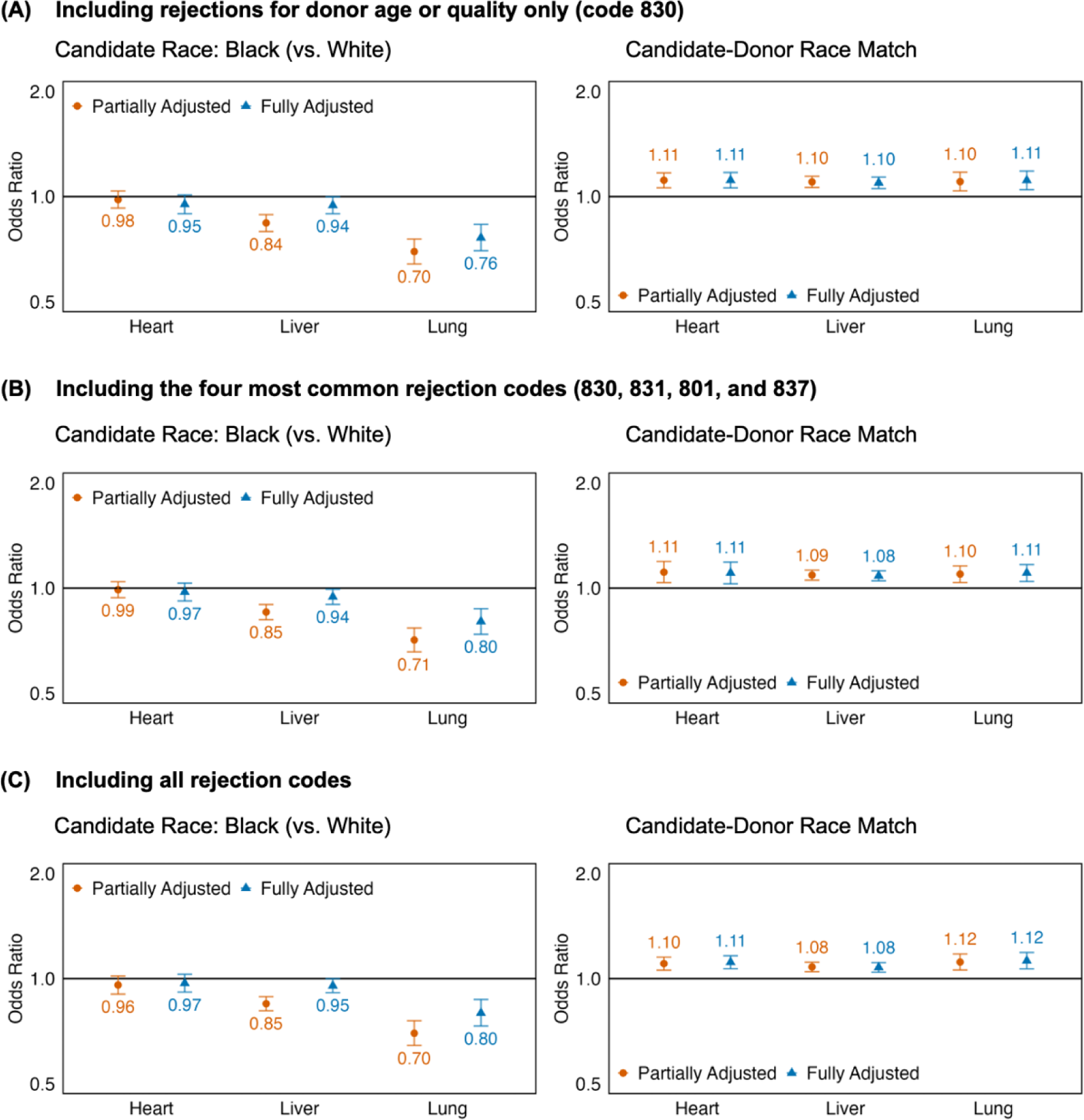
Robustness of main results to rejection code exclusion criterion. We change our cohort to include rejections for a broader or narrower set of reasons. In (A) we only include rejections attributed to code 830 (donor age or quality), the most frequently used reason. In (B), we expand the cohort to include rejections attributed to codes 830 (donor age or quality), 831 (donor size or weight), 801 (patient ill, unavailable, refused, or temporarily unsuitable), and 837 (organ-specific donor issue). In (C), we include all rejection codes (see Table S1 for a detailed list). We plot the odds ratios for the association of offer acceptance with Black candidate race (left) and donor-candidate race match (right). Error bars denote 95% confidence intervals, calculated using cluster-robust standard errors (clustering by candidate and donor). The results in (A), (B), and (C) mirror those presented in Fig. 4 in the main text.

**Figure S4.**
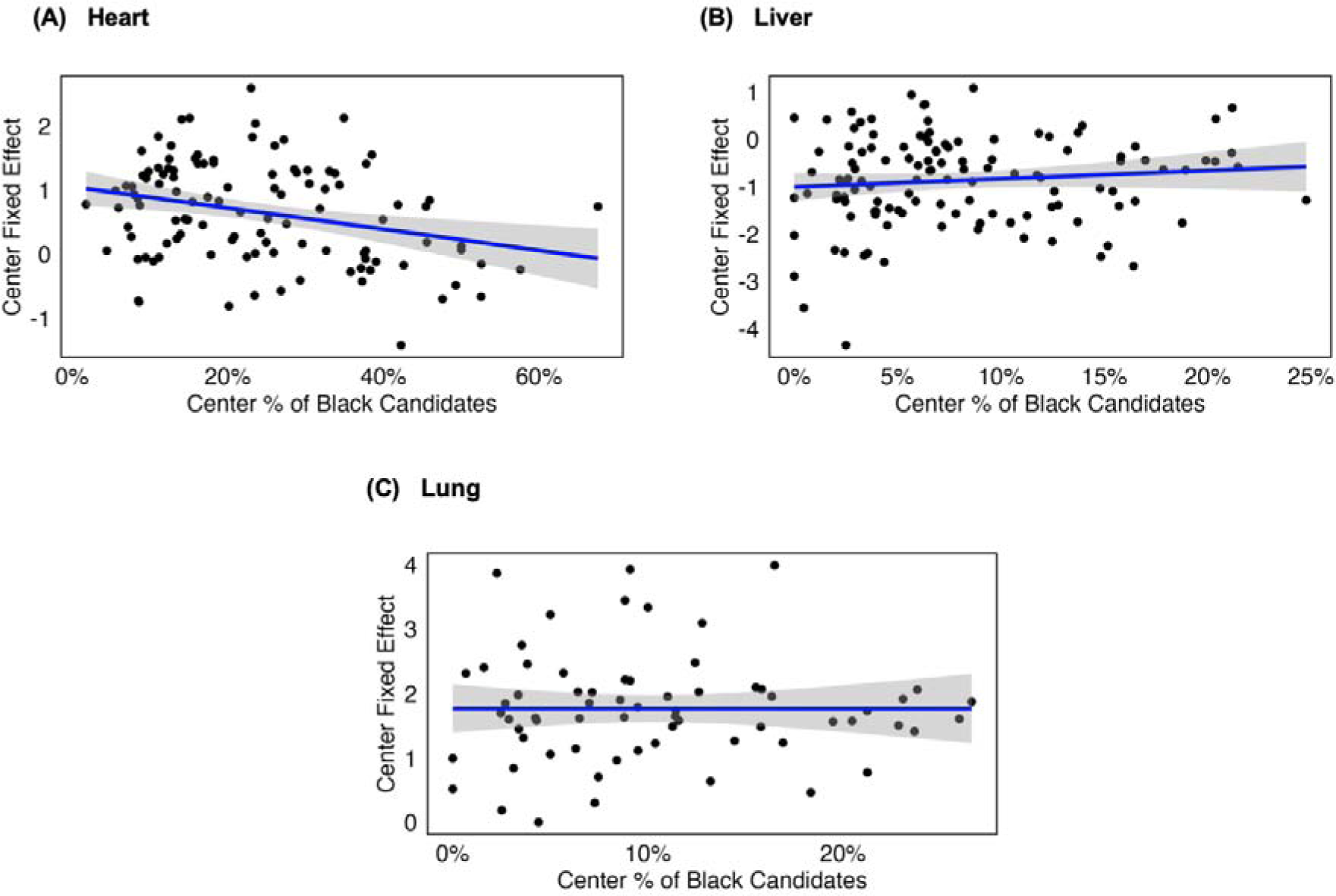
Transplant center mean acceptance propensity vs. proportion of Black patients. We plot the center-level fixed effects from the fully adjusted logistic regressions against the proportion of Black patients at each center for (A) hearts (B) livers and (C) lungs. The higher the fixed effect, the higher the center’s mean propensity to accept an offer. The lack of correlation for liver and lungs (p-value of non-zero effect: 0.25 for livers, 0.99 for lungs) suggests that Black patients are not overrepresented at centers with lower acceptance propensities. For hearts, there is evidence that Black patients are over-represented at centers with lower acceptance rates (regression coefficient -1.65, p-value 0.002).

**Figure S5.**
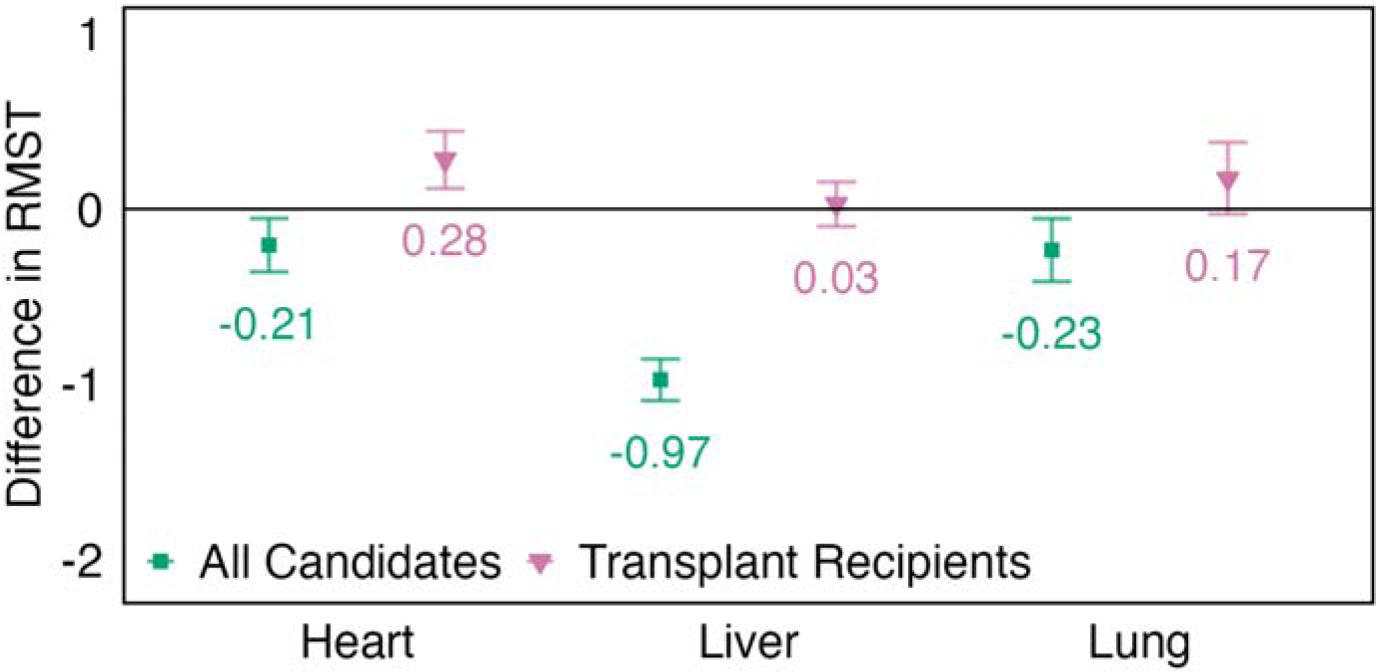
Analysis of restricted mean survival times. Differences in restricted mean survival time (RMST) for the effect of offer rejection on survival time. A negative difference indicates that patients with rejected offers have increased mortality risk than similarly ill patients with accepted offers. Error bars denote 95% confidence intervals.

### Supplementary Tables

**Table S1.** Summary of Offer Rejection Reasons.

| Code | Description | Heart | Liver | Lung |
| --- | --- | --- | --- | --- |
|  | Accepted | 31,631 (9%) | 73,286 (7%) | 23,479 (7%) |
| 830 | Donor age or quality | 137,128 (39%) | 633,283 (58%) | 150,071 (48%) |
| 831 | Donor size/weight | 97,083 (28%) | 100,806 (9%) | 61,841 (20%) |
| 801 | Patient ill, unavailable, refused, or temporarily unsuitable | 11,663 (3%) | 63,105 (6%) | 2,777 (1%) |
| 837 | Organ-specific donor issue | 17,298 (5%) | 35,342 (3%) | 22,712 (7%) |
| 898 | Other - Specify | 4,455 (1%) | 59,005 (5%) | 6,253 (2%) |
| 802 | Multiple organ transplant or different laterality is required | 5,543 (2%) | 26,608 (2%) | 23,639 (8%) |
| 803 | Patient txed, tx in progress, or other offer being considered | 7,307 (2%) | 26,239 (2%) | 3,297 (1%) |
| 824 | Distance to travel or ship | 6,950 (2%) | 11,055 (1%) | 2,465 (1%) |
| 833 | Donor social history | 7,286 (2%) | 6,483 (1%) | 4,706 (2%) |
| 835 | Organ Preservation | 442 (0%) | 13,923 (1%) | 609 (0%) |
| 813 | Unacceptable Antigens | 9,992 (3%) | 520 (0%) | 3,286 (1%) |
| 834 | Positive serological tests | 1,219 (0%) | 8,606 (1%) | 1,836 (1%) |
| 2000 | Miscellaneous bypass | 94 (0%) | 9,251 (1%) | 67 (0%) |
| 836 | Organ anatomical damage or defect | 1,280 (0%) | 4,196 (0%) | 1,119 (0%) |
| 812 | No serum for crossmatching | 4,254 (1%) | 22 (0%) | 918 (0%) |
| 810 | Positive crossmatch | 2,727 (1%) | 477 (0%) | 1,342 (0%) |
| 800 | Patient's condition improved, transplant not needed | 209 (0%) | 2,546 (0%) | 79 (0%) |
| 832 | Donor ABO | 10 (0%) | 2,333 (0%) | 56 (0%) |
| 842 | COVID-19: OPO or transplant hospital operational issue | 426 (0%) | 1,577 (0%) | 511 (0%) |
| 811 | Number of HLA mismatches unacceptable | 1,008 (0%) | 149 (0%) | 452 (0%) |
| 863 | Offer not made due to expedited placement attempt | - (0%) | 899 (0%) | - (0%) |
| 841 | COVID-19 donor-related reason | 288 (0%) | 658 (0%) | 210 (0%) |
| 825 | Operational – transplant center | 199 (0%) | 576 (0%) | 330 (0%) |
| 820 | Heavy workload | 286 (0%) | 443 (0%) | 312 (0%) |
| 823 | Surgeon unavailable | 229 (0%) | 274 (0%) | 276 (0%) |
| 860 | Medical urgency of another potential recipient | 289 (0%) | 127 (0%) | 377 (0%) |
| 822 | Exceeded one hour response time | 71 (0%) | 256 (0%) | 124 (0%) |
| 840 | COVID-19: candidate-related reason | 147 (0%) | 273 (0%) | 16 (0%) |
| 815 | High CPRA | 292 (0%) | 34 (0%) | 49 (0%) |
| 881 | Maximum Omm offer limits exceeded | 57 (0%) | 159 (0%) | 34 (0%) |
| 2001 | Cold ischemic time | - (0%) | 127 (0%) | - (0%) |
| Total |  | 349,863 | 1,082,638 | 313,243 |

**Table S2.** Candidate race before and after applying exclusion criteria.

|  | Heart |  | Liver |  | Lung |  |
| --- | --- | --- | --- | --- | --- | --- |
|  | Before<br>Exclusion<br>(N=46,836) | After<br>Exclusion<br>(N=32,268) | Before<br>Exclusion<br>(N=147,605) | After<br>Exclusion<br>(N=102,823) | Before<br>Exclusion<br>(N=31,054) | After<br>Exclusion<br>(N=25,780) |
| **Race** |  |  |  |  |  |  |
| White | 29,279<br>(62.5%) | 20,553<br>(63.7%) | 101,742<br>(68.9%) | 71,947<br>(70.0%) | 24,419<br>(78.6%) | 20,358<br>(79.0%) |
| Black | 10,769<br>(23.0%) | 7,669 (23.8%) | 12,987 (8.8%) | 8,820 (8.6%) | 3,027 (9.7%) | 2,563 (9.9%) |
| Hispanic | 4,609 (9.8%) | 2,629 (8.1%) | 23,932 (16.2%) | 16,006<br>(15.6%) | 2,635 (8.5%) | 2,066 (8.0%) |
| Other | 2,179 (4.7%) | 1,417 (4.4%) | 8,944 (6.1%) | 6,050 (5.9%) | 973 (3.1%) | 793 (3.1%) |

**Table S3.** Comparison of rejecting and accepting groups in the survival analysis cohort. N is the number of donor organs; for each organ, we include two candidates, one who accepted the offer and one who rejected the offer. The rejecting candidate was ranked immediately above the accepting candidate. We describe the differences in blood type and allocation score between the accepting and rejecting candidate for each organ, finding that the two groups are well-matched.

|  | Heart | Liver | Lung |
| --- | --- | --- | --- |
| **Characteristic** | **N = 6,773** | **N = 13,685** | **N = 5,592** |
| Identical blood type<sup>1</sup> | 5,290 (78%) | 12,602 (92%) | 5,211 (93%) |
| Identical priority status<sup>1</sup> | 6,346 (94%) |  |  |
| Difference in MELD<sup>2</sup> |  | 0.0 (-2.0, 0.0) |  |
| Difference in LAS<sup>2</sup> |  |  | -1 (-3, 0) |

<sup>1</sup> n (%)
<sup>2</sup> Median (IQR)

**Table S4.**
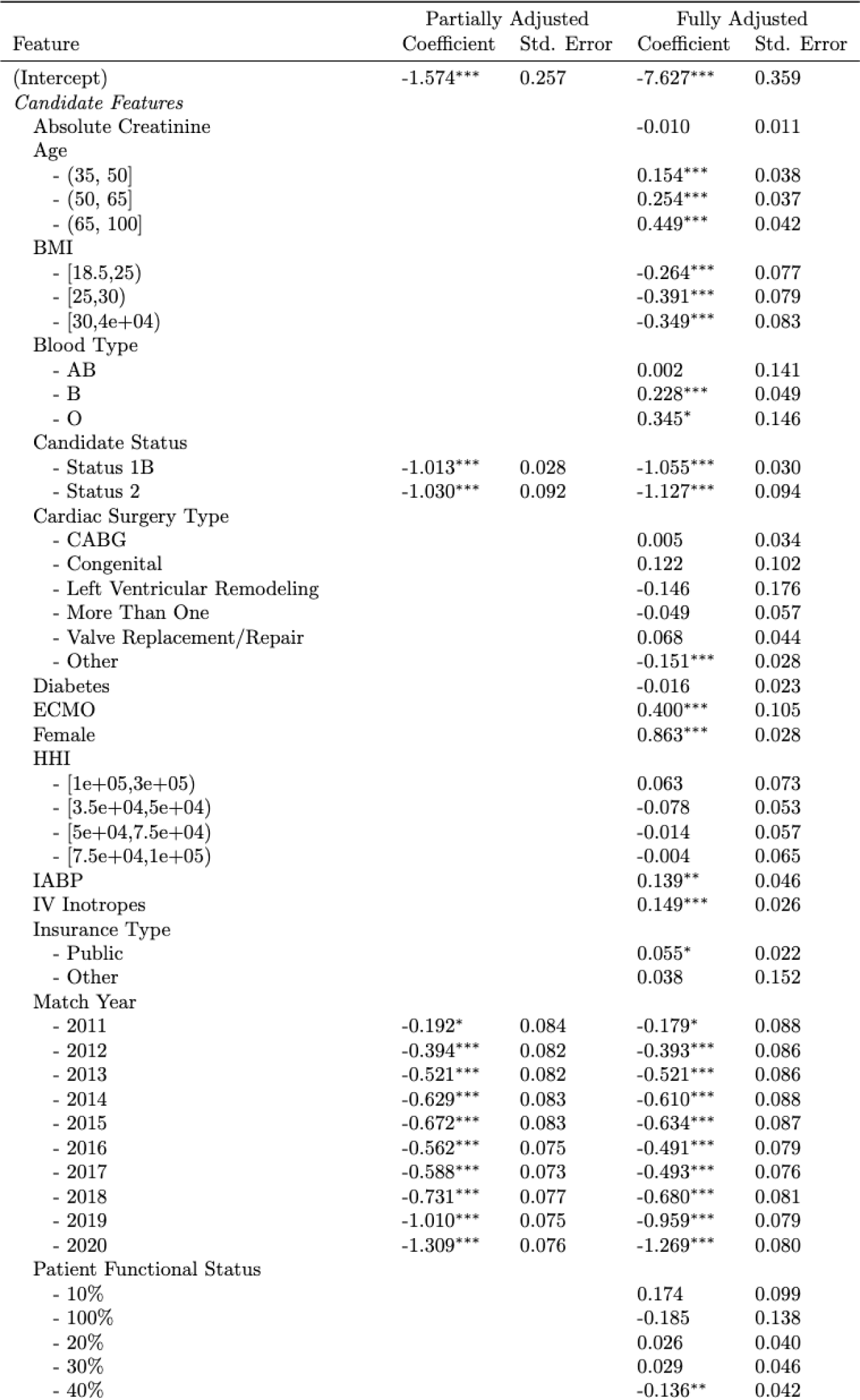
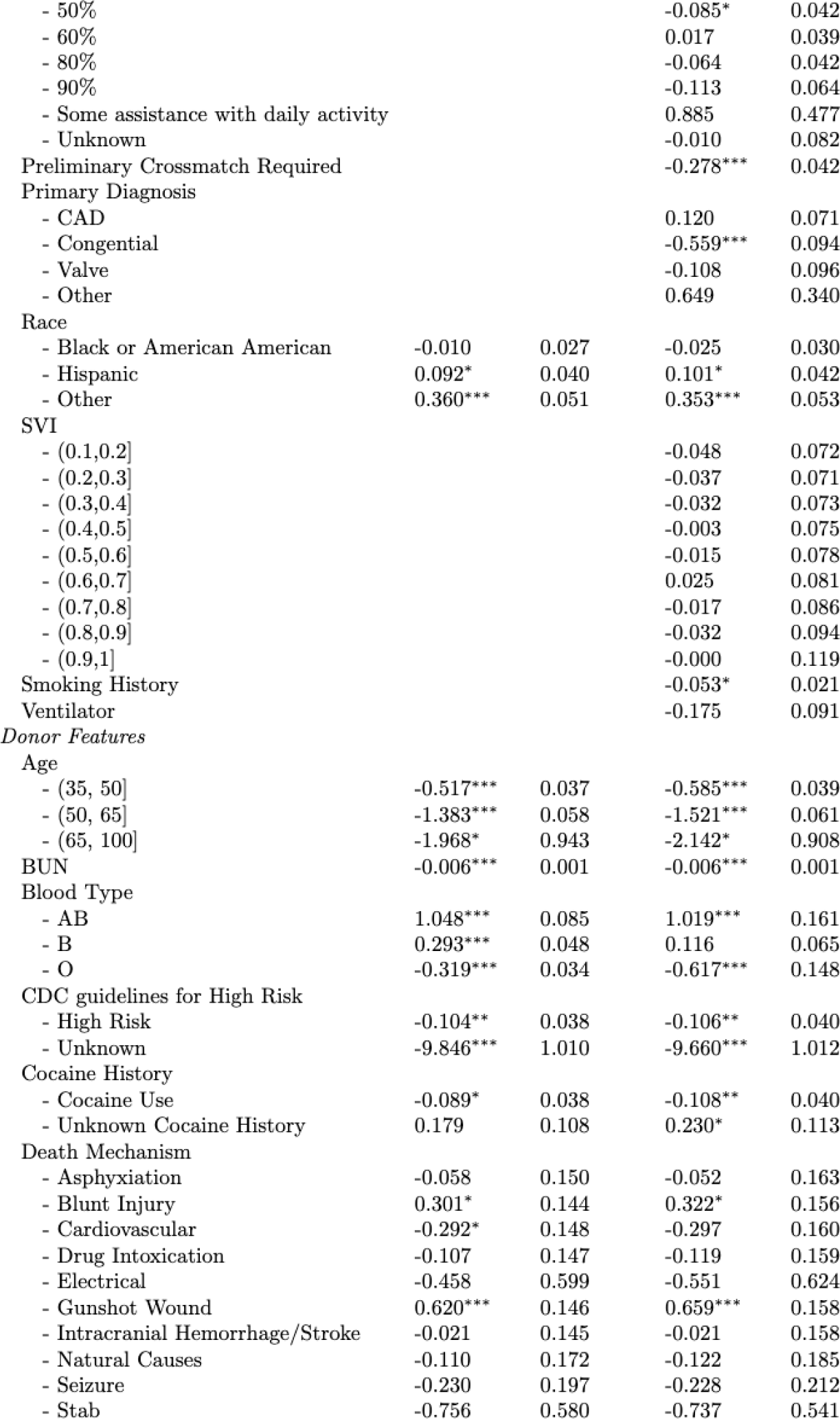
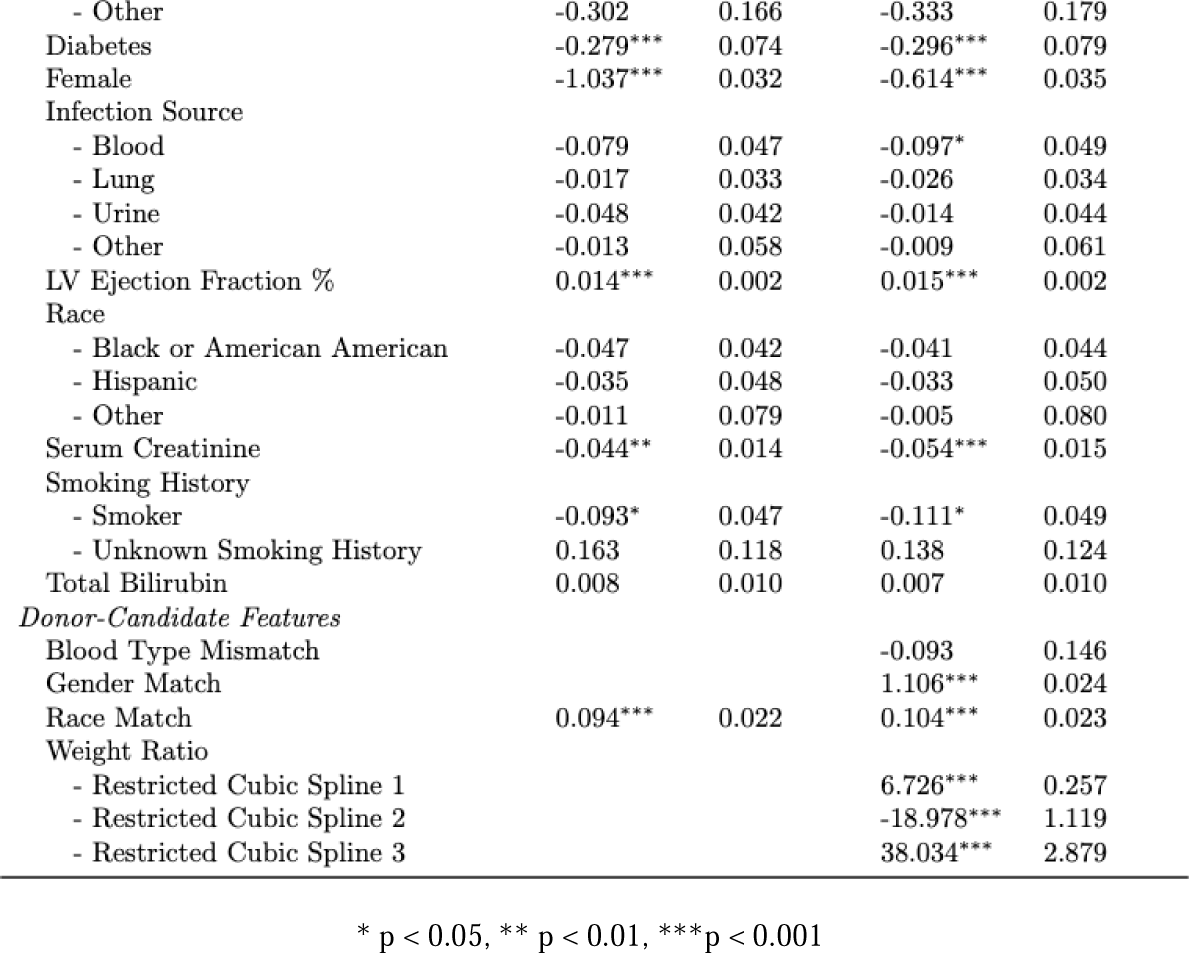
Coefficients from logistic regression for heart transplant acceptance.

**Table S5.**
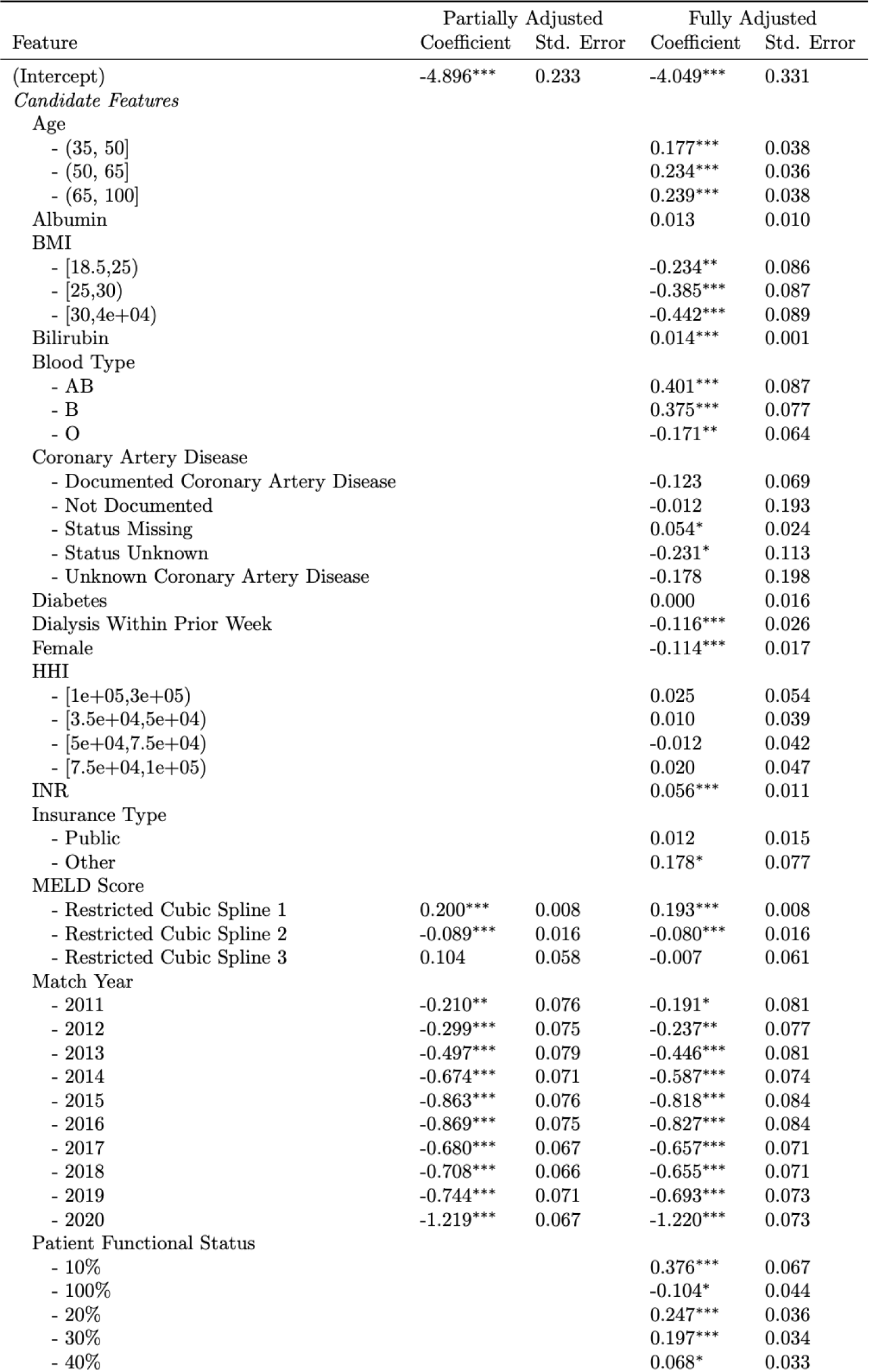
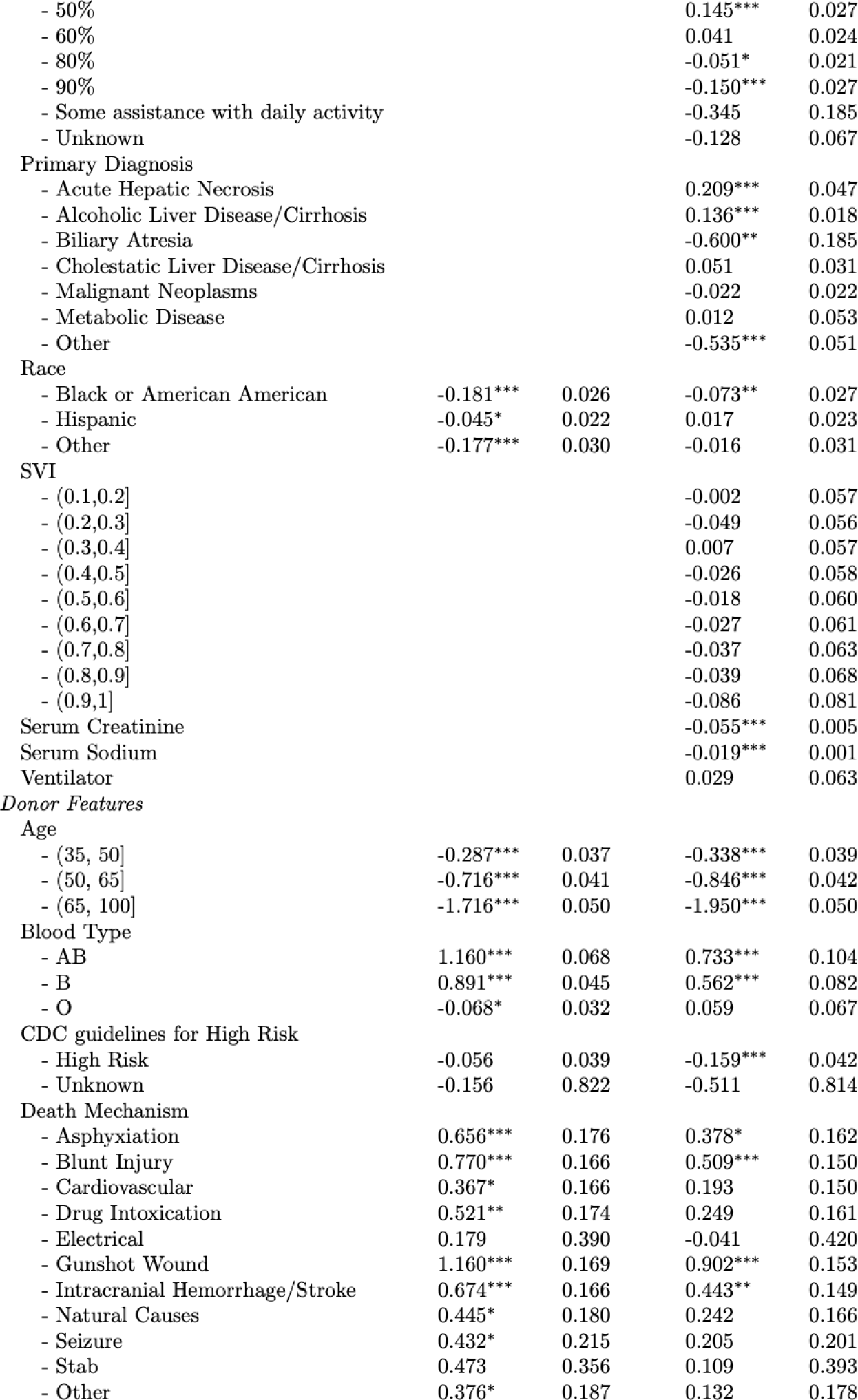
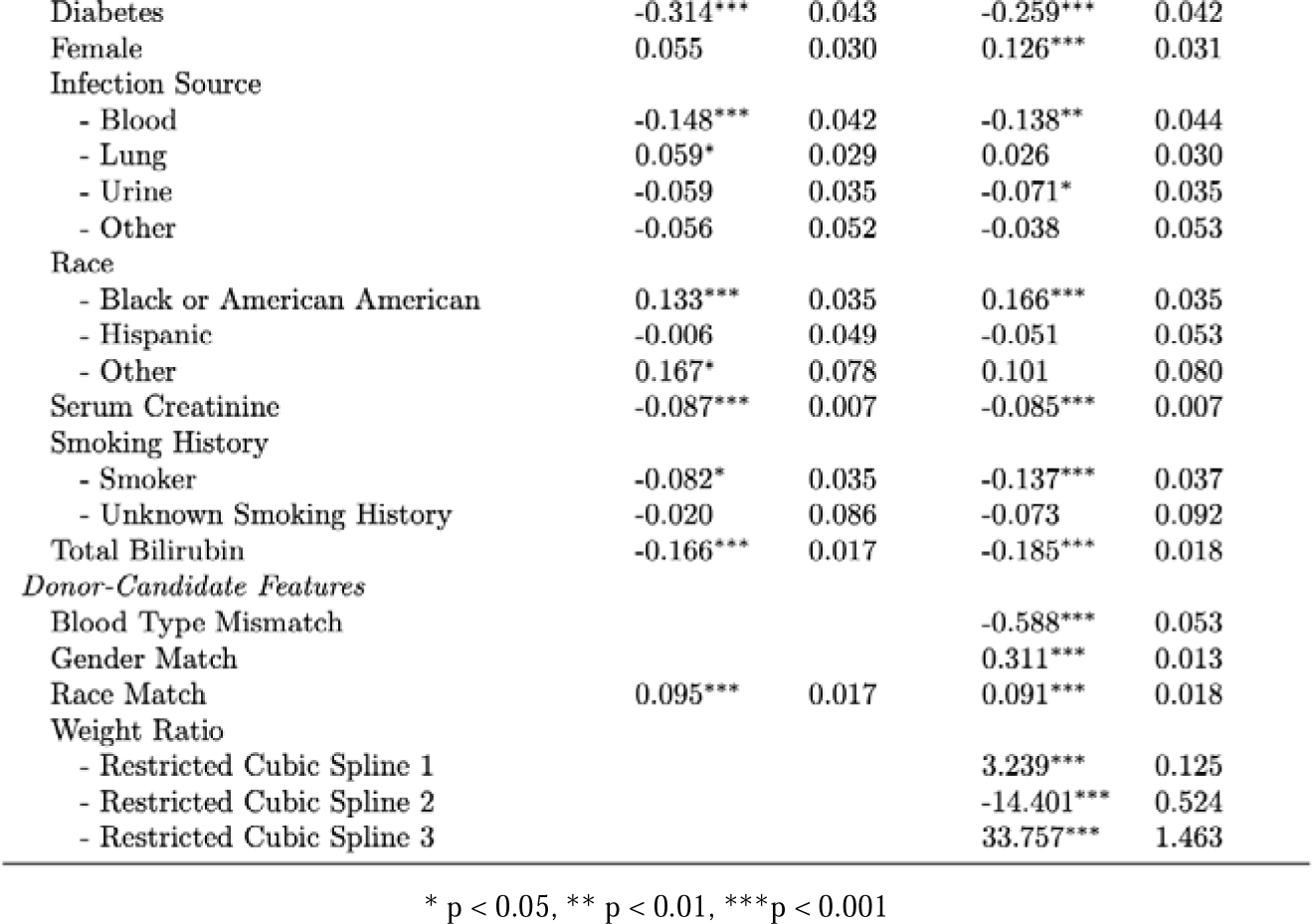
Coefficients from logistic regression for liver transplant acceptance.

| Feature | Partially Adjusted |  | Fully Adjusted |  |
| --- | --- | --- | --- | --- |
|  | Coefficient | Std. Error | Coefficient | Std. Error |
| (Intercept) | -4.896*** | 0.233 | -4.049*** | 0.331 |
| *Candidate Features* |  |  |  |  |
| Age |  |  |  |  |
| - (35, 50] |  |  | 0.177*** | 0.038 |
| - (50, 65] |  |  | 0.234*** | 0.036 |
| - (65, 100] |  |  | 0.239*** | 0.038 |
| Albumin |  |  | 0.013 | 0.010 |
| BMI |  |  |  |  |
| - [18.5,25) |  |  | -0.234** | 0.086 |
| - [25,30) |  |  | -0.385*** | 0.087 |
| - [30,4e+04) |  |  | -0.442*** | 0.089 |
| Bilirubin |  |  | 0.014*** | 0.001 |
| Blood Type |  |  |  |  |
| - AB |  |  | 0.401*** | 0.087 |
| - B |  |  | 0.375*** | 0.077 |
| - O |  |  | -0.171** | 0.064 |
| Coronary Artery Disease |  |  |  |  |
| - Documented Coronary Artery Disease |  |  | -0.123 | 0.069 |
| - Not Documented |  |  | -0.012 | 0.193 |
| - Status Missing |  |  | 0.054* | 0.024 |
| - Status Unknown |  |  | -0.231* | 0.113 |
| - Unknown Coronary Artery Disease |  |  | -0.178 | 0.198 |
| Diabetes |  |  | 0.000 | 0.016 |
| Dialysis Within Prior Week |  |  | -0.116*** | 0.026 |
| Female |  |  | -0.114*** | 0.017 |
| HHI |  |  |  |  |
| - [1e+05,3e+05) |  |  | 0.025 | 0.054 |
| - [3.5e+04,5e+04) |  |  | 0.010 | 0.039 |
| - [5e+04,7.5e+04) |  |  | -0.012 | 0.042 |
| - [7.5e+04,1e+05) |  |  | 0.020 | 0.047 |
| INR |  |  | 0.056*** | 0.011 |
| Insurance Type |  |  |  |  |
| - Public |  |  | 0.012 | 0.015 |
| - Other |  |  | 0.178* | 0.077 |
| MELD Score |  |  |  |  |
| - Restricted Cubic Spline 1 | 0.200*** | 0.008 | 0.193*** | 0.008 |
| - Restricted Cubic Spline 2 | -0.089*** | 0.016 | -0.080*** | 0.016 |
| - Restricted Cubic Spline 3 | 0.104 | 0.058 | -0.007 | 0.061 |
| Match Year |  |  |  |  |
| - 2011 | -0.210** | 0.076 | -0.191* | 0.081 |
| - 2012 | -0.299*** | 0.075 | -0.237** | 0.077 |
| - 2013 | -0.497*** | 0.079 | -0.446*** | 0.081 |
| - 2014 | -0.674*** | 0.071 | -0.587*** | 0.074 |
| - 2015 | -0.863*** | 0.076 | -0.818*** | 0.084 |
| - 2016 | -0.869*** | 0.075 | -0.827*** | 0.084 |
| - 2017 | -0.680*** | 0.067 | -0.657*** | 0.071 |
| - 2018 | -0.708*** | 0.066 | -0.655*** | 0.071 |
| - 2019 | -0.744*** | 0.071 | -0.693*** | 0.073 |
| - 2020 | -1.219*** | 0.067 | -1.220*** | 0.073 |
| Patient Functional Status |  |  |  |  |
| - 10% |  |  | 0.376*** | 0.067 |
| - 100% |  |  | -0.104* | 0.044 |
| - 20% |  |  | 0.247*** | 0.036 |
| - 30% |  |  | 0.197*** | 0.034 |
| - 40% |  |  | 0.068* | 0.033 |
|  | Coefficient | Std. Error | Coefficient | Std. Error |
| - 50% |  |  | 0.145*** | 0.027 |
| - 60% |  |  | 0.041 | 0.024 |
| - 80% |  |  | -0.051* | 0.021 |
| - 90% |  |  | -0.150*** | 0.027 |
| - Some assistance with daily activity |  |  | -0.345 | 0.185 |
| - Unknown |  |  | -0.128 | 0.067 |
| Primary Diagnosis |  |  |  |  |
| - Acute Hepatic Necrosis |  |  | 0.209*** | 0.047 |
| - Alcoholic Liver Disease/Cirrhosis |  |  | 0.136*** | 0.018 |
| - Biliary Atresia |  |  | -0.600** | 0.185 |
| - Cholestatic Liver Disease/Cirrhosis |  |  | 0.051 | 0.031 |
| - Malignant Neoplasms |  |  | -0.022 | 0.022 |
| - Metabolic Disease |  |  | 0.012 | 0.053 |
| - Other |  |  | -0.535*** | 0.051 |
| Race |  |  |  |  |
| - Black or American American | -0.181*** | 0.026 | -0.073** | 0.027 |
| - Hispanic | -0.045* | 0.022 | 0.017 | 0.023 |
| - Other | -0.177*** | 0.030 | -0.016 | 0.031 |
| SVI |  |  |  |  |
| - (0.1,0.2] |  |  | -0.002 | 0.057 |
| - (0.2,0.3] |  |  | -0.049 | 0.056 |
| - (0.3,0.4] |  |  | 0.007 | 0.057 |
| - (0.4,0.5] |  |  | -0.026 | 0.058 |
| - (0.5,0.6] |  |  | -0.018 | 0.060 |
| - (0.6,0.7] |  |  | -0.027 | 0.061 |
| - (0.7,0.8] |  |  | -0.037 | 0.063 |
| - (0.8,0.9] |  |  | -0.039 | 0.068 |
| - (0.9,1] |  |  | -0.086 | 0.081 |
| Serum Creatinine |  |  | -0.055*** | 0.005 |
| Serum Sodium |  |  | -0.019*** | 0.001 |
| Ventilator |  |  | 0.029 | 0.063 |
| *Donor Features* |  |  |  |  |
| Age |  |  |  |  |
| - (35, 50] | -0.287*** | 0.037 | -0.338*** | 0.039 |
| - (50, 65] | -0.716*** | 0.041 | -0.846*** | 0.042 |
| - (65, 100] | -1.716*** | 0.050 | -1.950*** | 0.050 |
| Blood Type |  |  |  |  |
| - AB | 1.160*** | 0.068 | 0.733*** | 0.104 |
| - B | 0.891*** | 0.045 | 0.562*** | 0.082 |
| - O | -0.068* | 0.032 | 0.059 | 0.067 |
| CDC guidelines for High Risk |  |  |  |  |
| - High Risk | -0.056 | 0.039 | -0.159*** | 0.042 |
| - Unknown | -0.156 | 0.822 | -0.511 | 0.814 |
| Death Mechanism |  |  |  |  |
| - Asphyxiation | 0.656*** | 0.176 | 0.378* | 0.162 |
| - Blunt Injury | 0.770*** | 0.166 | 0.509*** | 0.150 |
| - Cardiovascular | 0.367* | 0.166 | 0.193 | 0.150 |
| - Drug Intoxication | 0.521** | 0.174 | 0.249 | 0.161 |
| - Electrical | 0.179 | 0.390 | -0.041 | 0.420 |
| - Gunshot Wound | 1.160*** | 0.169 | 0.902*** | 0.153 |
| - Intracranial Hemorrhage/Stroke | 0.674*** | 0.166 | 0.443** | 0.149 |
| - Natural Causes | 0.445* | 0.180 | 0.242 | 0.166 |
| - Seizure | 0.432* | 0.215 | 0.205 | 0.201 |
| - Stab | 0.473 | 0.356 | 0.109 | 0.393 |
| - Other | 0.376* | 0.187 | 0.132 | 0.178 |
|  | Coefficient | Std. Error | Coefficient | Std. Error |
| Diabetes | -0.314*** | 0.043 | -0.259*** | 0.042 |
| Female | 0.055 | 0.030 | 0.126*** | 0.031 |
| Infection Source |  |  |  |  |
| - Blood | -0.148*** | 0.042 | -0.138** | 0.044 |
| - Lung | 0.059* | 0.029 | 0.026 | 0.030 |
| - Urine | -0.059 | 0.035 | -0.071* | 0.035 |
| - Other | -0.056 | 0.052 | -0.038 | 0.053 |
| Race |  |  |  |  |
| - Black or American American | 0.133*** | 0.035 | 0.166*** | 0.035 |
| - Hispanic | -0.006 | 0.049 | -0.051 | 0.053 |
| - Other | 0.167* | 0.078 | 0.101 | 0.080 |
| Serum Creatinine | -0.087*** | 0.007 | -0.085*** | 0.007 |
| Smoking History |  |  |  |  |
| - Smoker | -0.082* | 0.035 | -0.137*** | 0.037 |
| - Unknown Smoking History | -0.020 | 0.086 | -0.073 | 0.092 |
| Total Bilirubin | -0.166*** | 0.017 | -0.185*** | 0.018 |
| *Donor-Candidate Features* |  |  |  |  |
| Blood Type Mismatch |  |  | -0.588*** | 0.053 |
| Gender Match |  |  | 0.311*** | 0.013 |
| Race Match | 0.095*** | 0.017 | 0.091*** | 0.018 |
| Weight Ratio |  |  |  |  |
| - Restricted Cubic Spline 1 |  |  | 3.239*** | 0.125 |
| - Restricted Cubic Spline 2 |  |  | -14.401*** | 0.524 |
| - Restricted Cubic Spline 3 |  |  | 33.757*** | 1.463 |

\* p < 0.05, \*\* p < 0.01, \*\*\*p < 0.001

**Table S6.**
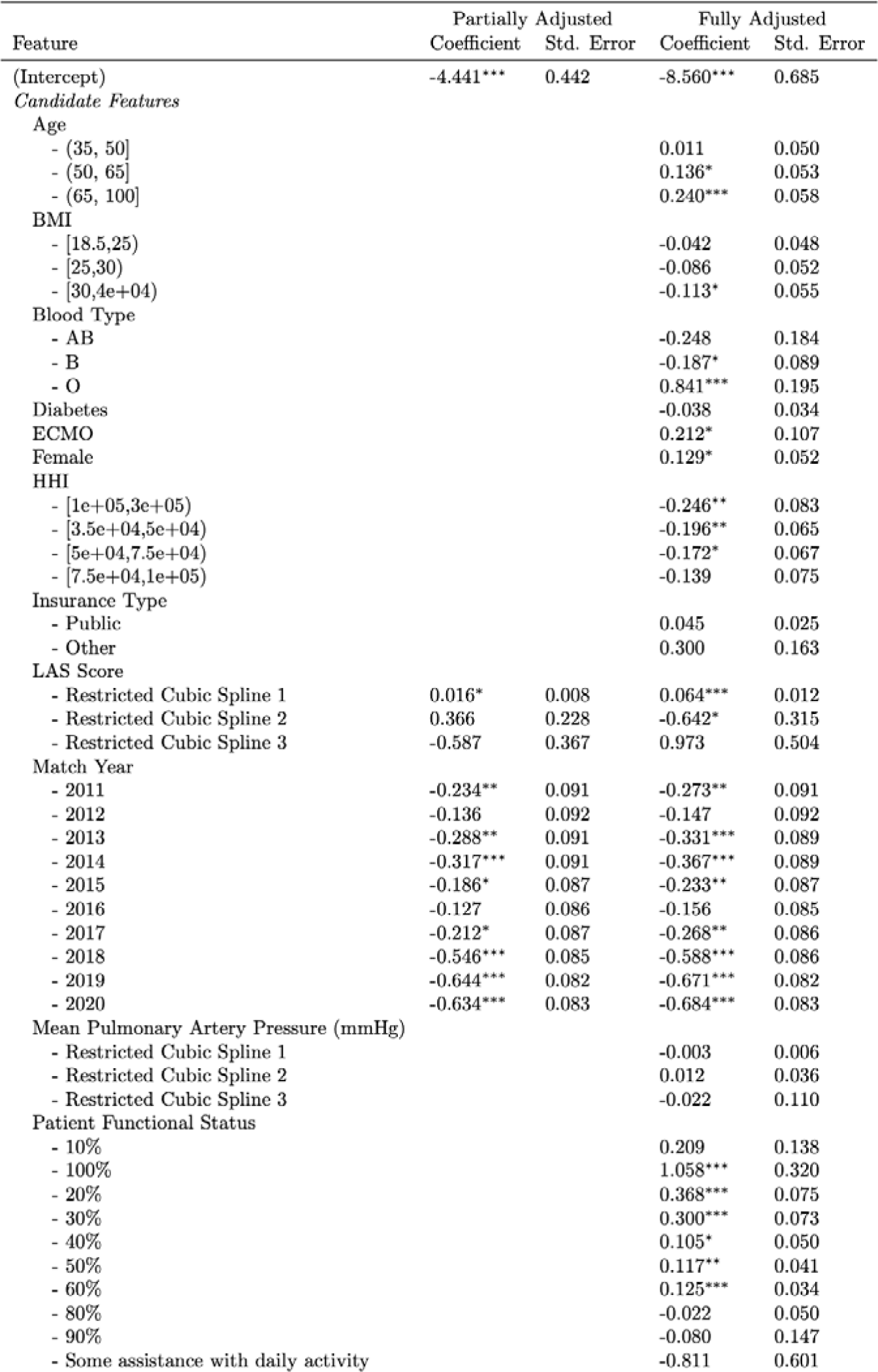
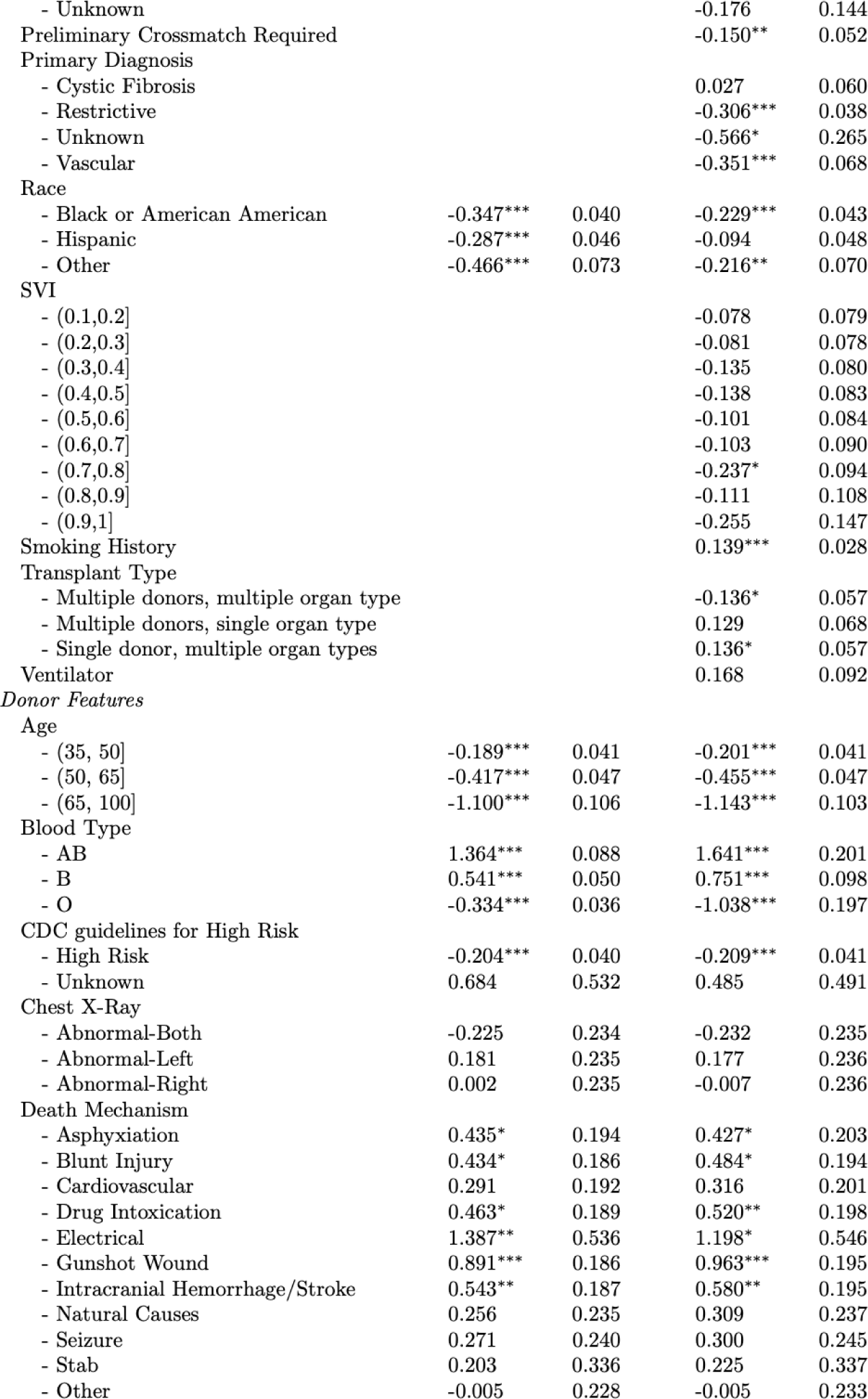
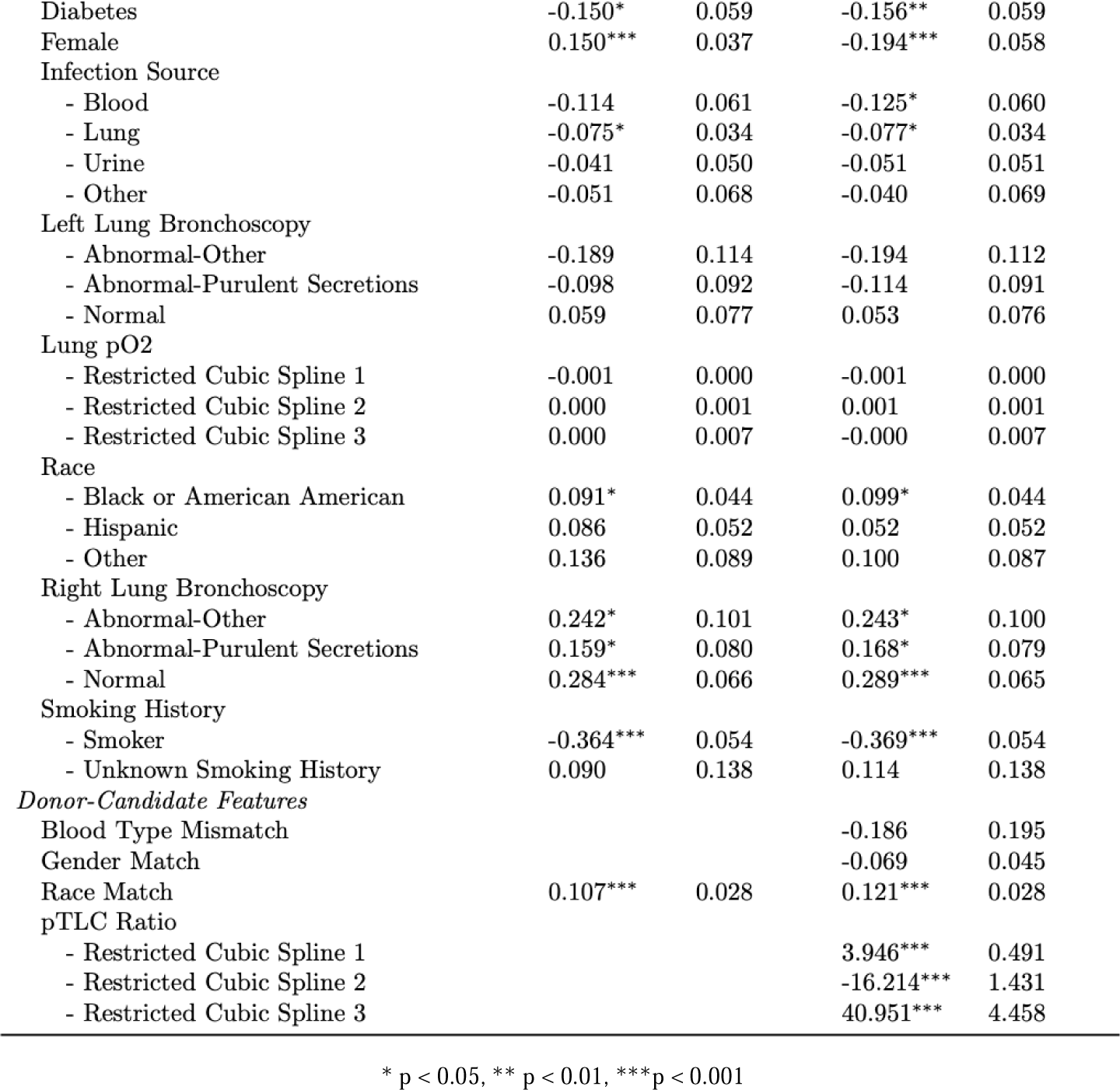
Coefficients from logistic regression for lung transplant acceptance.

| Feature | Partially Adjusted |  | Fully Adjusted |  |
| --- | --- | --- | --- | --- |
|  | Coefficient | Std. Error | Coefficient | Std. Error |
| (Intercept) | -4.441*** | 0.442 | -8.560*** | 0.685 |
| *Candidate Features* |  |  |  |  |
| Age |  |  |  |  |
| - (35, 50] |  |  | 0.011 | 0.050 |
| - (50, 65] |  |  | 0.136* | 0.053 |
| - (65, 100] |  |  | 0.240*** | 0.058 |
| BMI |  |  |  |  |
| - [18.5,25) |  |  | -0.042 | 0.048 |
| - [25,30) |  |  | -0.086 | 0.052 |
| - [30,4e+04) |  |  | -0.113* | 0.055 |
| Blood Type |  |  |  |  |
| - AB |  |  | -0.248 | 0.184 |
| - B |  |  | -0.187* | 0.089 |
| - O |  |  | 0.841*** | 0.195 |
| Diabetes |  |  | -0.038 | 0.034 |
| ECMO |  |  | 0.212* | 0.107 |
| Female |  |  | 0.129* | 0.052 |
| HHI |  |  |  |  |
| - [1e+05,3e+05) |  |  | -0.246** | 0.083 |
| - [3.5e+04,5e+04) |  |  | -0.196** | 0.065 |
| - [5e+04,7.5e+04) |  |  | -0.172* | 0.067 |
| - [7.5e+04,1e+05) |  |  | -0.139 | 0.075 |
| Insurance Type |  |  |  |  |
| - Public |  |  | 0.045 | 0.025 |
| - Other |  |  | 0.300 | 0.163 |
| LAS Score |  |  |  |  |
| - Restricted Cubic Spline 1 | 0.016* | 0.008 | 0.064*** | 0.012 |
| - Restricted Cubic Spline 2 | 0.366 | 0.228 | -0.642* | 0.315 |
| - Restricted Cubic Spline 3 | -0.587 | 0.367 | 0.973 | 0.504 |
| Match Year |  |  |  |  |
| - 2011 | -0.234** | 0.091 | -0.273** | 0.091 |
| - 2012 | -0.136 | 0.092 | -0.147 | 0.092 |
| - 2013 | -0.288** | 0.091 | -0.331*** | 0.089 |
| - 2014 | -0.317*** | 0.091 | -0.367*** | 0.089 |
| - 2015 | -0.186* | 0.087 | -0.233** | 0.087 |
| - 2016 | -0.127 | 0.086 | -0.156 | 0.085 |
| - 2017 | -0.212* | 0.087 | -0.268** | 0.086 |
| - 2018 | -0.546*** | 0.085 | -0.588*** | 0.086 |
| - 2019 | -0.644*** | 0.082 | -0.671*** | 0.082 |
| - 2020 | -0.634*** | 0.083 | -0.684*** | 0.083 |
| Mean Pulmonary Artery Pressure (mmHg) |  |  |  |  |
| - Restricted Cubic Spline 1 |  |  | -0.003 | 0.006 |
| - Restricted Cubic Spline 2 |  |  | 0.012 | 0.036 |
| - Restricted Cubic Spline 3 |  |  | -0.022 | 0.110 |
| Patient Functional Status |  |  |  |  |
| - 10% |  |  | 0.209 | 0.138 |
| - 100% |  |  | 1.058*** | 0.320 |
| - 20% |  |  | 0.368*** | 0.075 |
| - 30% |  |  | 0.300*** | 0.073 |
| - 40% |  |  | 0.105* | 0.050 |
| - 50% |  |  | 0.117** | 0.041 |
| - 60% |  |  | 0.125*** | 0.034 |
| - 80% |  |  | -0.022 | 0.050 |
| - 90% |  |  | -0.080 | 0.147 |
| - Some assistance with daily activity |  |  | -0.811 | 0.601 |
|  | Coefficient | Std. Error | Coefficient | Std. Error |
| - Unknown |  |  | -0.176 | 0.144 |
| Preliminary Crossmatch Required |  |  | -0.150** | 0.052 |
| Primary Diagnosis |  |  |  |  |
| - Cystic Fibrosis |  |  | 0.027 | 0.060 |
| - Restrictive |  |  | -0.306*** | 0.038 |
| - Unknown |  |  | -0.566* | 0.265 |
| - Vascular |  |  | -0.351*** | 0.068 |
| Race |  |  |  |  |
| - Black or American American | -0.347*** | 0.040 | -0.229*** | 0.043 |
| - Hispanic | -0.287*** | 0.046 | -0.094 | 0.048 |
| - Other | -0.466*** | 0.073 | -0.216** | 0.070 |
| SVI |  |  |  |  |
| - (0.1,0.2] |  |  | -0.078 | 0.079 |
| - (0.2,0.3] |  |  | -0.081 | 0.078 |
| - (0.3,0.4] |  |  | -0.135 | 0.080 |
| - (0.4,0.5] |  |  | -0.138 | 0.083 |
| - (0.5,0.6] |  |  | -0.101 | 0.084 |
| - (0.6,0.7] |  |  | -0.103 | 0.090 |
| - (0.7,0.8] |  |  | -0.237* | 0.094 |
| - (0.8,0.9] |  |  | -0.111 | 0.108 |
| - (0.9,1] |  |  | -0.255 | 0.147 |
| Smoking History |  |  | 0.139*** | 0.028 |
| Transplant Type |  |  |  |  |
| - Multiple donors, multiple organ type |  |  | -0.136* | 0.057 |
| - Multiple donors, single organ type |  |  | 0.129 | 0.068 |
| - Single donor, multiple organ types |  |  | 0.136* | 0.057 |
| Ventilator |  |  | 0.168 | 0.092 |
| *Donor Features* |  |  |  |  |
| Age |  |  |  |  |
| - (35, 50] | -0.189*** | 0.041 | -0.201*** | 0.041 |
| - (50, 65] | -0.417*** | 0.047 | -0.455*** | 0.047 |
| - (65, 100] | -1.100*** | 0.106 | -1.143*** | 0.103 |
| Blood Type |  |  |  |  |
| - AB | 1.364*** | 0.088 | 1.641*** | 0.201 |
| - B | 0.541*** | 0.050 | 0.751*** | 0.098 |
| - O | -0.334*** | 0.036 | -1.038*** | 0.197 |
| CDC guidelines for High Risk |  |  |  |  |
| - High Risk | -0.204*** | 0.040 | -0.209*** | 0.041 |
| - Unknown | 0.684 | 0.532 | 0.485 | 0.491 |
| Chest X-Ray |  |  |  |  |
| - Abnormal-Both | -0.225 | 0.234 | -0.232 | 0.235 |
| - Abnormal-Left | 0.181 | 0.235 | 0.177 | 0.236 |
| - Abnormal-Right | 0.002 | 0.235 | -0.007 | 0.236 |
| Death Mechanism |  |  |  |  |
| - Asphyxiation | 0.435* | 0.194 | 0.427* | 0.203 |
| - Blunt Injury | 0.434* | 0.186 | 0.484* | 0.194 |
| - Cardiovascular | 0.291 | 0.192 | 0.316 | 0.201 |
| - Drug Intoxication | 0.463* | 0.189 | 0.520** | 0.198 |
| - Electrical | 1.387** | 0.536 | 1.198* | 0.546 |
| - Gunshot Wound | 0.891*** | 0.186 | 0.963*** | 0.195 |
| - Intracranial Hemorrhage/Stroke | 0.543** | 0.187 | 0.580** | 0.195 |
| - Natural Causes | 0.256 | 0.235 | 0.309 | 0.237 |
| - Seizure | 0.271 | 0.240 | 0.300 | 0.245 |
| - Stab | 0.203 | 0.336 | 0.225 | 0.337 |
| - Other | -0.005 | 0.228 | -0.005 | 0.233 |
|  | Coefficient | Std. Error | Coefficient | Std. Error |
| Diabetes | -0.150* | 0.059 | -0.156** | 0.059 |
| Female | 0.150*** | 0.037 | -0.194*** | 0.058 |
| Infection Source |  |  |  |  |
| - Blood | -0.114 | 0.061 | -0.125* | 0.060 |
| - Lung | -0.075* | 0.034 | -0.077* | 0.034 |
| - Urine | -0.041 | 0.050 | -0.051 | 0.051 |
| - Other | -0.051 | 0.068 | -0.040 | 0.069 |
| Left Lung Bronchoscopy |  |  |  |  |
| - Abnormal-Other | -0.189 | 0.114 | -0.194 | 0.112 |
| - Abnormal-Purulent Secretions | -0.098 | 0.092 | -0.114 | 0.091 |
| - Normal | 0.059 | 0.077 | 0.053 | 0.076 |
| Lung pO2 |  |  |  |  |
| - Restricted Cubic Spline 1 | -0.001 | 0.000 | -0.001 | 0.000 |
| - Restricted Cubic Spline 2 | 0.000 | 0.001 | 0.001 | 0.001 |
| - Restricted Cubic Spline 3 | 0.000 | 0.007 | -0.000 | 0.007 |
| Race |  |  |  |  |
| - Black or American American | 0.091* | 0.044 | 0.099* | 0.044 |
| - Hispanic | 0.086 | 0.052 | 0.052 | 0.052 |
| - Other | 0.136 | 0.089 | 0.100 | 0.087 |
| Right Lung Bronchoscopy |  |  |  |  |
| - Abnormal-Other | 0.242* | 0.101 | 0.243* | 0.100 |
| - Abnormal-Purulent Secretions | 0.159* | 0.080 | 0.168* | 0.079 |
| - Normal | 0.284*** | 0.066 | 0.289*** | 0.065 |
| Smoking History |  |  |  |  |
| - Smoker | -0.364*** | 0.054 | -0.369*** | 0.054 |
| - Unknown Smoking History | 0.090 | 0.138 | 0.114 | 0.138 |
| *Donor-Candidate Features* |  |  |  |  |
| Blood Type Mismatch |  |  | -0.186 | 0.195 |
| Gender Match |  |  | -0.069 | 0.045 |
| Race Match | 0.107*** | 0.028 | 0.121*** | 0.028 |
| pTLC Ratio |  |  |  |  |
| - Restricted Cubic Spline 1 |  |  | 3.946*** | 0.491 |
| - Restricted Cubic Spline 2 |  |  | -16.214*** | 1.431 |
| - Restricted Cubic Spline 3 |  |  | 40.951*** | 4.458 |

\* p < 0.05, \*\* p < 0.01, \*\*\*p < 0.001

**Table S7.** Interaction of race and transplant center size. We present results from fully adjusted models of offer acceptance that include an interaction term between transplant center size and our two variables of interest: candidate race and donor-candidate race match. For each organ, we rank centers by the number of offers received (in our cohort), defining large centers as those in the upper quartile. The table displays the estimated odds ratios for the baseline terms (corresponding to non-large centers) and the interaction terms (corresponding to multiplicative change in odds ratios from non-large centers to large centers). None of the interaction coefficients for Black race are significant at a 0.05 level; this indicates that the observed associations between Black race and offer acceptance are neither significantly smaller nor larger at large centers (compared to small centers). There is some evidence that the association between race match and acceptance is reduced at large centers for livers, though not for hearts and lungs.

|  | Heart | Liver | Lung |
| --- | --- | --- | --- |
| **Baseline odds ratios** |  |  |  |
| Black race (vs. White Race) | 0.97 (0.90, 1.05) | 0.91 (0.85, 0.98) | 0.81 (0.73, 0.91) |
| Donor-candidate race match | 1.12 (1.06, 1.19) | 1.13 (1.08, 1.19) | 1.14 (1.06, 1.22) |
| **Interaction odds-ratios** |  |  |  |
| Black race : Large center | 1.01 (0.90, 1.12) | 1.04 (0.94, 1.15) | 0.96 (0.82, 1.11) |
| Race match : Large center | 0.97 (0.89, 1.05) | 0.94 (0.88, 0.99) | 0.99 (0.91, 1.08) |

**Table S8.**
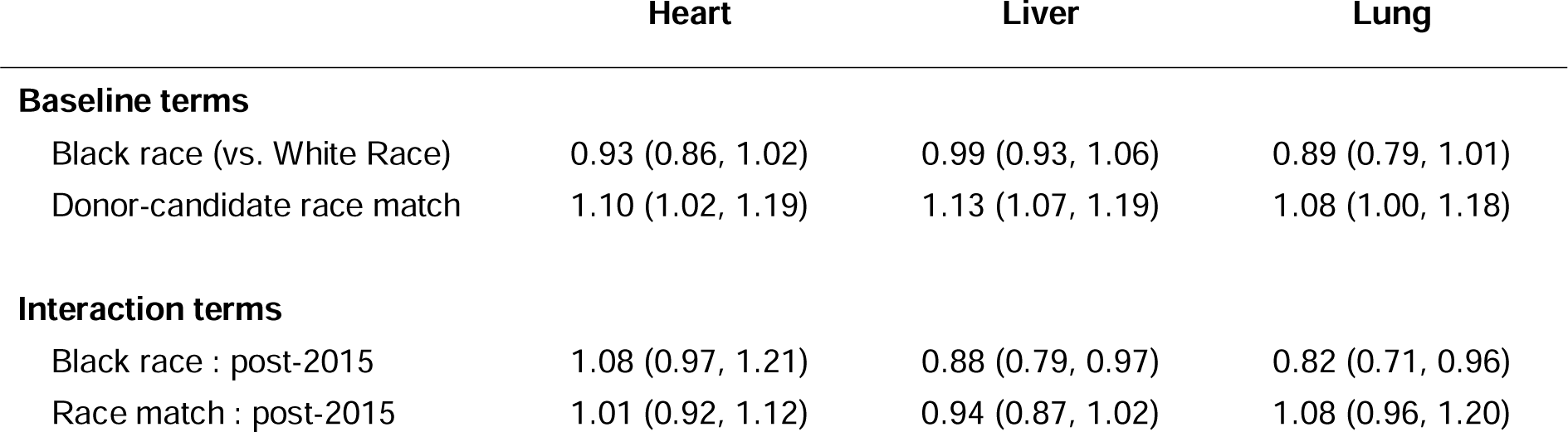
Interaction of race and time. We present results from fully adjusted models of offer acceptance that include an interaction term between time and our two variables of interest: candidate race and donor-candidate race match. We consider two time periods: the first six years of our study period (i.e., 2010-2015) and the last five years (i.e., 2016-2020). The table displays the estimated odds ratios for the baseline terms (corresponding to the 2010-2015 period) and the interaction terms (corresponding to multiplicative change in odds ratios from the first period to the second period). The interaction odds ratios for Black race are significantly smaller than 1 for livers and lungs, indicating that the negative association between Black race and offer acceptance has increased over time.

## Notes

### Competing Interest Statement

The authors have declared no competing interest.

### Author Declarations

This study was granted full approval following expedited review by the Committee on the Use of Humans as Experimental Subjects, the Institutional Review Board for the Massachusetts Institute of Technology.

### Summary of Updates

Updated acknowledgements to correct a typo

